# Identifying Multi-omics Signatures that characterize Responders to Plant-based Dietary Interventions

**DOI:** 10.1101/2025.11.06.25339667

**Authors:** Alina Schieren, Hanna Huber, Carolina Alvarez-Garavito, Aakash Mantri, Waldemar Seel, Ramona Dolscheid-Pommerich, Martin Coenen, Matthias Schmid, Bolette Hartmann, Jens J. Holst, Mohamed Yaghmour, Christoph Thiele, Markus M. Nöthen, Jan Hasenauer, Peter Stehle, Marie-Christine Simon

## Abstract

Metabolic responses to dietary interventions often show high inter-individual variations, but the factors explaining these variations as well as underlying mechanisms and their interrelation remain largely unknown. Knowing which factors are relevant for the individual response to a dietary intervention and to which extent, is essential to provide targeted dietary recommendations. Therefore, in persons at increased cardiometabolic risk, a 6-week RCT comparing the individual effects of three dietary patterns was conducted. We observed highly individualized effects in the cholesterol-lowering ability of the dietary patterns, which seem mainly related to differences in the lipid and metabolic profile of the participants in addition to diet-specific characteristics, while the gut microbiome and the polygenetic risk for hyperlipidemia modulate the effects. Moreover, the individual response to the interventions might be predicted by machine-learning models providing multi-omics signatures that could be applied as biomarkers for stratification of persons to specific dietary interventions for the optimization of personalized dietary approaches.

## INTRODUCTION

Modifying diet is a key element of strategies targeting overweight, obesity and cardiovascular disease (CVD) risk^1–3^. In recent years, the focus in nutrition and health research shifted from a generalized to a more personalized approach based on the observation that food components, foods and diets may lead to differential metabolic responses in humans, and a concept of personalized nutrition has developed. The metabolic and genetic profile as well as the gut-microbiome might be important influencing factors determining how a person responds to a diet modification^4–11^. However, multi-omics approaches, combining several data layers, as well as lipidomics data, providing additional information on cardiometabolic risk^12–15^; have not been regularly implemented in dietary intervention studies. Especially, predicting the success of a dietary intervention based on these data, by for example applying machine-learning models, is a new approach.

Especially in people at high CVD risk, improving lipid(-omic) and glycemic profile is of high relevance and greatly impacts disease progression and risk^16–21^. In particular, low-density lipoprotein cholesterol (LDL-C) is a well-established CVD risk marker and a causal factor in the pathophysiology of atherosclerosis. Therefore, lowering LDL-C levels is a key target of prevention strategies with great risk-reduction potential^22–25^. Besides pharmaceutical approaches like glucagon-like peptide 1 (GLP-1) analogues or statins^26–28^, adapting plant-based dietary patterns, focusing primarily on foods from plant origin while minimizing or eliminating animal products, is a promising strategy for CVD risk reduction as they have been associated with reductions of body weight, CVD risk factors and mortality^1–3,29,30^. Among the beneficial plant-based dietary patterns, especially an ovo-lacto Vegetarian diet as well as a ‘Nordic Diet’, comprising foods traditionally available in the Scandinavian region such as berries, wholegrain oats and rye, root vegetables, and fatty sea fish have demonstrated health-enhancing effects and great potential for CVD prevention^31–35^. In contrast, a western-type dietary pattern has been shown to contribute to the development of metabolic diseases and obesity^36^.

We hypothesized that the response, as defined by the reduction in LDL-C, to a dietary intervention in adults at elevated cardiometabolic risk, is highly individual and dependent on the baseline biological profile. Therefore, the aim of this study was to investigate features characterizing responders (RS) and non-responders (NRS) to specific plant-based dietary interventions, applying single-and multi-omics approaches with data comprising 16S-derived metagenomics, fecal short-chain fatty acids, lipidomic profile, metabolic and anthropometric parameters and genetic risk scores for hypercholesterinemia and triacylglycerides (TAG). Specifically, a Nordic and a Vegetarian dietary pattern were examined in comparison to the participant’s habitual western-type dietary pattern. Thus, we performed model-based feature selection and tailored univariate testing of the identified features. Moreover, we applied machine-learning models predicting the success of the dietary interventions, further elucidating underlying factors influencing the individual differences in responses. If the success of dietary interventions can be predicted from deep characterization of an individual’s metabolic, genetic, lipidomic and microbial profile at baseline, using the identified features as biomarkers may facilitate a more targeted and personalized approach for dietary interventions.

## RESULTS

This study is a randomized, controlled intervention trial in parallel design, comprising two study phases of 6-weeks duration each (**Fig. 1 A**). Of the 120 recruited participants of the 1^st^ study phase, 112 completed the study. In the 2^nd^ study phase, 26 of 27 randomized participants completed the protocol **(Fig. S1).**

**Figure 1:**
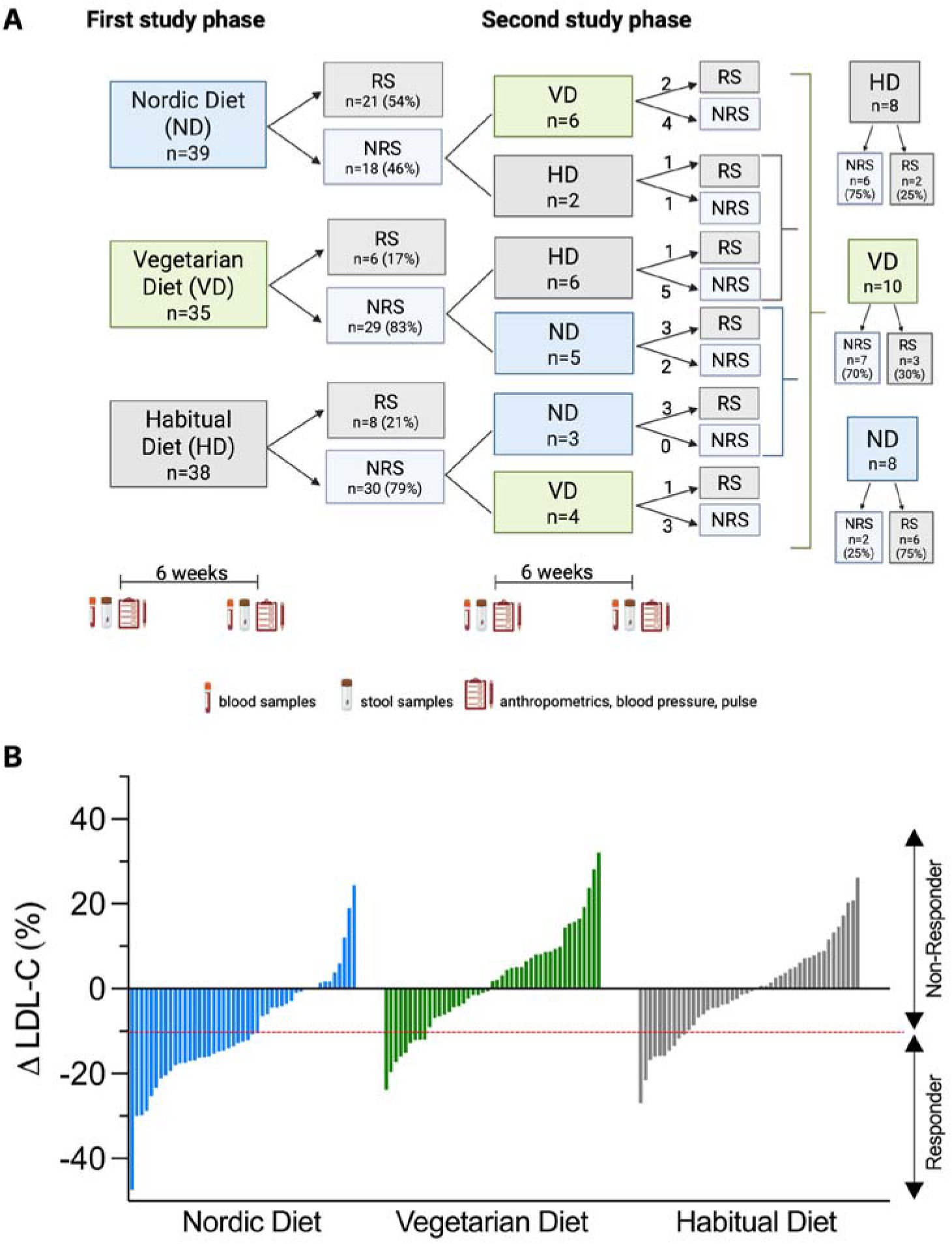
Study design and individual response to the interventions. **A.** Study design consisting of two independent study phases of six weeks duration, each. Participants were randomized to one of three diet groups: ND, VD or HD (control group). After each study phase, participants were classified into RS or NRS according to their change in LDL-C. In the second study phase, only a subgroup of NRS of the first study participated and were randomized in a cross-over design to one of the remaining two diets. The figure was created using www.biorender.com. B. Bar graphs showing the individual relative changes in LDL-C from baseline to endline in ND, VD and HD group for each participant. The red line shows the cut-off (-10% relative change) selected for classification of RS and NRS. *HD, habitual diet; LDL-C, low-density lipoprotein cholesterol; ND, Nordic diet; NRS, Non-Responders, RS, Responders; VD, Vegetarian diet*

The baseline characteristics of the study cohort are reported in **Table 1**. The BMI, waist-to-height ratio, fat mass, blood pressure and total cholesterol (TC) levels were elevated in all participants, reflecting the high cardiometabolic risk of the cohort.

**Table 1:** Baseline characteristics of the study participants in the different diet groups.

| Parameters | All diets combined<br>(n=138) | ND<br>(n=47) | VD<br>(n=45) | HD<br>(n=46) |
| --- | --- | --- | --- | --- |
| Sex (women/men, %) | 59/41 | 66/34 | 60/40 | 52/48 |
| Age (years) | 59.4±0.6 | 60.3±1 | 59.2±1.1 | 58.8±1.1 |
| BMI (kg/m<sup>2</sup>) | 31±0.3 | 30.7±0.5 | 31.3±0.6 | 31.1±0.5 |
| Waist-to-height Ratio | 0.62±0 | 0.62±0 | 0.63±0 | 0.61±0 |
| Fat mass (%) | 41.3±0.7 | 41.3±1.2 | 42.6±1.2 | 40±1.3 |
| SBP (mmHg) | 136±1 | 136±2 | 138±3 | 134±2 |
| DBP (mmHg) | 87±1 | 88±2 | 88±2 | 86±2 |
| Pulse (bpm) | 68±1 | 67±2 | 68±2 | 68±2 |
| RMR (kcal/day) | 1713±25 | 1683±39 | 1673±45 | 1784±43 |
| TC (mmol/l) | 5.6±0.1 | 6±0.2 | 5.6±0.1 | 5.3±0.2 |
| LDL-C (mmol/l) | 3.8±0.1 | 4.1±0.1 | 3.8±0.1 | 3.5±0.1 |
| HDL-C (mmol/l) | 1.48±0.03 | 1.52±0.05 | 1.42±0.05 | 1.49±0.06 |

Mean + SEM. BMI: body mass Index. DBP: diastolic blood pressure; HD: Habitual Diet; HDL-C: high density lipoprotein cholesterol; LDL-C: low density lipoprotein cholesterol; ND: Nordic Diet; NRS: Non-Responders; RMR: resting metabolic rate; RS: Responders; SBP: systolic blood pressure; TC: total cholesterol; VD: Vegetarian Diet.

The isocaloric intervention diets led to a change in LDL-C values ranging from-2.4 mmol/l (-47%) up to +1.17 mmol/l (+32%) **(Fig. 1 B)**. Participants with a reduction in LDL-C of at least 10% after the 6-weeks intervention were categorized as RS, while all remaining participants were defined as NRS. In total 33% of the participants were classified as RS, 57% in the Nordic Diet (ND) group, 20% in the Vegetarian Diet (VD) group and 22% in the control group (Habitual Diet (HD)) **(Fig. 1 A)**. The change in LDL-C after the interventions showed a marked inter-individual variation, with the greatest effects induced by the ND. In total, 39% of the women and 25% of the men classified as RS.

### RS are generally characterized by a less favorable baseline clinical lipid and glucose metabolism

To characterize RS and NRS in depth, feature selection based on single-and multi-omics approaches was performed using ‘sparse partial least squares-discriminant analysis (sPLS-DA)’ for single-omics and supervised N-integration sPLS-DA with DIABLO (Data Integration Analysis for Biomarker discovery using Latent variable approaches for Omics studies) for multi-omics models in each diet group. Model performance was evaluated using AUC and balanced error rate (BER) derived from 5-fold cross-validation. RS and NRS in general (combining all diet groups) were distinguished best by the following baseline features: *Parabacteroides*, the ND and VD group, LDL-C, TC, high-density lipoprotein cholesterol (HDL-C), ether-linked lyso-phosphatidylcholine (LPC-O) (16:0) and polygenetic risk score (PGS) PGS_LDL-C_ in the DIABLO model (AUC=0.652, *p=*0.028, BER_centroids_=0.31) (**Table S1**). Tailored univariate analysis of the top features confirmed that RS of all diets combined were characterized by significantly higher initial levels of TC, LDL-C (both *p_un_*_adj_<0.001) and HDL-C (*p*_unadj_=0.036) and slightly higher levels of LPC-O (16:0) (*p*_unadj_=0.009) compared to NRS (**Fig. 2 A, Table S2**). Univariate Pearson correlation analysis showed that baseline LDL-C levels correlate positively with TC, LPC-O (16:0), PGS_LDL-C_ and inversely with *Parabacteroides* (**Table S3**). The metabolic dataset had the highest contribution to classifying RS and NRS as indicated by the block weight (0.44) (**Fig. 2 B**, **Table S1**). Also within the single-omics analyses with sPLS-DA, the metabolic dataset classified RS and NRS best (**Fig. 2 C**, **Table S1**). The sPLS-DA models (**Fig. 2 D**) confirmed the importance of the top features of the DIABLO model and revealed additional relevant microbes and lipid species which were distinctive of RS and NRS (**Fig. 2 A**, **Table S1**).

**Figure 2:**
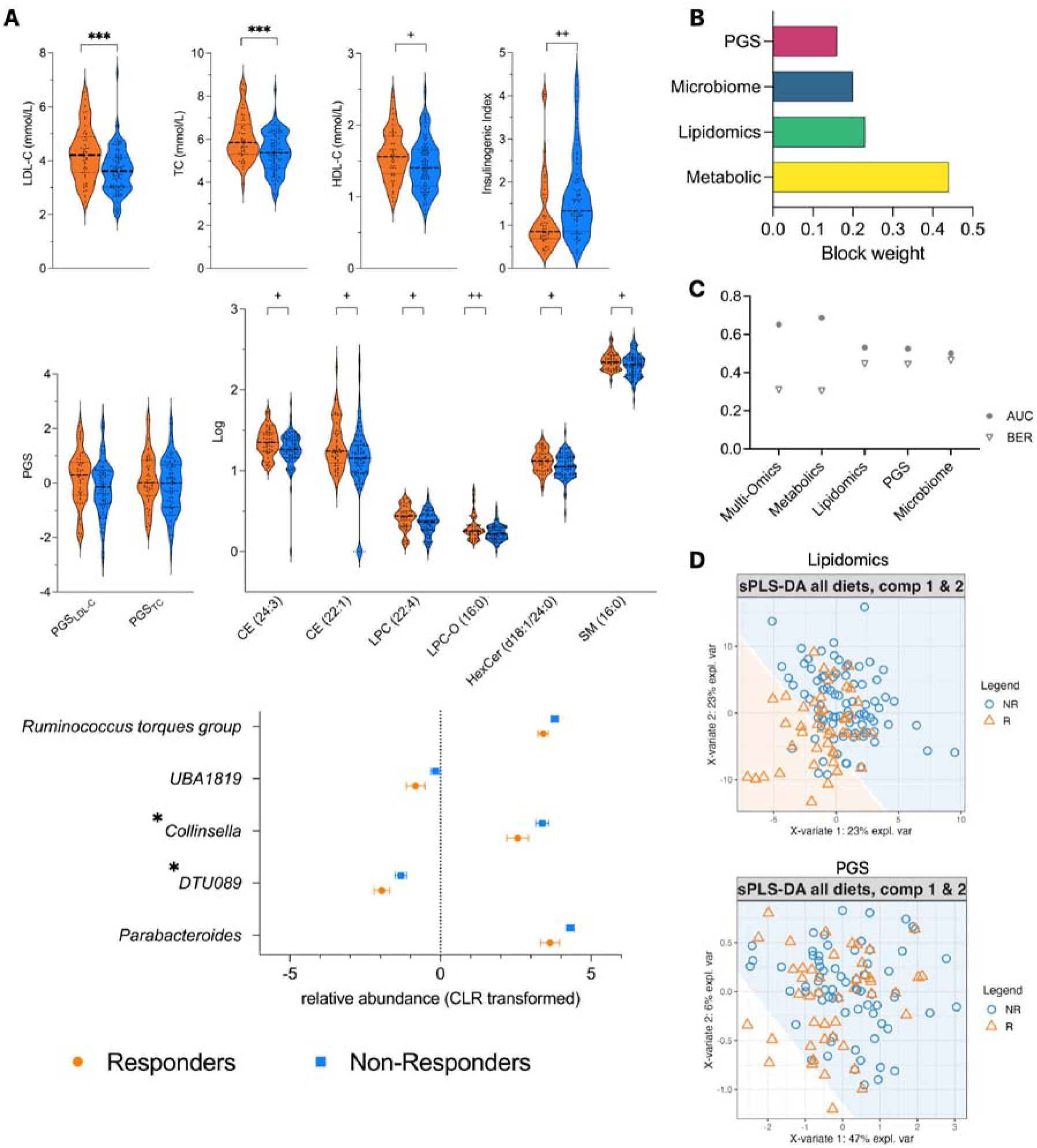
Models distinguishing RS and NRS in all diets combined at baseline. A: Violin and forest plots show levels or relative abundances + SEM (raw values for metabolic dataset, log transformed data for lipidomic dataset, CLR transformed data for microbiome dataset) of the top selected features from DIABLO, sPLS-DA and XGBoost models distinguishing RS (orange, *n*=46) and NRS (blue, *n*=92) from all diets combined at baseline. For visualization purposes, outliers of the insulinogenic index are not shown. Between-group differences were analyzed using Student’s t-test for metabolic data, linear regression or zero-inflated gaussian models for lipidomic data and Wilcoxon test for microbiome data. \*\*\**p*_adj_<0.001, \**p*_adj_<0.05, ++*p*_unadj_<0.01, +*p_unadj_*<0.05. For *Collinsella*, *DTU089* and SM (16:0) results were not confirmed by sensitivity analysis. B: Bar plot show contribution (block weight) of the four omics datasets to each component of the DIABLO model. C: Plot shows the performance, measured by AUC and BER of the multi-omics (DIABLO) and single-omics (sPLS-DA) analyses distinguishing RS and NRS from all diets combined at baseline. D: Sample plots from sPLS-DA include a prediction background to show the class assignment to RS (orange) or NRS (blue) for new data points based on their values on the first two latent components for the single-omics datasets. For lipidomic and PGS datasets, only components 1 and 2 are shown. No plot is shown for metabolic and microbiome datasets, as they have only 1 component. See also Table 1, S1, S2, S3, S4. *AUC, area under the curve; BER, balanced error rate; CE, cholesteryl esters; DAG, diacylglycerides; DIABLO, Data Integration Analysis for Biomarker discovery using Latent variable approaches for Omics studies; HD, Habitual Diet; HDL-C, high-density lipoprotein cholesterol; HexCer, hexosylceramides; LDL-C, low-density lipoprotein cholesterol; LPC, lysophosphatidylcholines; LPC-O, ether-linked lysophosphatidylcholines; ND, Nordic Diet; NRS, Non-Responders; p*_adj_, *adjusted p-value; PGS, polygenetic risk scores;* p_unadj_, *unadjusted p-value; RS, Responders; SEM, standard error of mean; SM, sphingomyelins; sPLS-DA, sparse Partial Least Squares - Discriminant Analysis; TAG, triacylglycerides; TC, total cholesterol; VD, Vegetarian Diet*.

Next, a machine learning algorithm was trained to predict the response to the diets based on baseline multi-and single-omics datasets with XGBoost^37^. For all diets combined, the XGBoost multi-omics prediction model had a sensitivity of 0.686, specificity of 0.444, an F1-score of 0.694 and an Area Under the Receiver Operating Characteristic Curve (AUC-ROC) of 0.591 (**Table S4**). The strongest predictors were the ND, insulinogenic index, LDL-C, SM (16:0) and LPC-O (16:0). Univariate analysis confirmed a lower insulinogenic index in RS (*p*_unadj_=0.007) (**Fig. 2 A**). Among the top 30 features were also TC and PGS_LDL-C_ confirming these features from the DIABLO model (**Fig. S2 A**). Of the single-omics datasets, the PGS led to the best prediction results which even exceeded the predictive accuracy of the multi-omics model with PGS_LDL-C_ as most important feature (**Table S4**).

These results indicate that irrespective of the specific dietary intervention, participants with a less favorable baseline clinical lipid and glucose metabolism profile are more likely to benefit. In addition, the abundance of specific gut microbes such as *Parabacteroides* are a relevant factor for the response, despite comparable diversity measures. The higher number of RS after following the ND compared to the VD was reflected in the ND variable being the top predictor variable in the XGBoost model and strongest contributor to the metabolic data component in the DIABLO model, indicating that the specific diet is a crucial factor for the individual response.

### A less favorable postprandial glucose metabolism at baseline characterizes RS to the Nordic Diet

For ND, the most important baseline features were *Ruminococcaceae UCG-005,* insulinogenic index, OGIS and disposition index, total fasting and postprandial TAG, PGS_TAG_, PGS_LDL-C_, PGS_TC_ and the lipid species diacylglycerol (DAG) (36:2) in the DIABLO multi-omics model (AUC=0.706, *p*=0.039, BER_max_=0.374) (**Table S1**). Tailored univariate analyses confirmed a significant difference in disposition index, OGIS (both *p*_unadj_≤0.01) and DAG (36:2) (p_unadj_=0.02) with lower values in RS of the ND compared to NRS and a significantly higher PGS score for LDL-C (p_unadj_=0.038) (**Fig. 3 A, Table S2**). There was a mild inverse correlation between *Ruminococcaceae UCG-005* and insulinogenic and disposition index, total fasting and postprandial TAG and DAG (36:2) in univariate correlation analysis (**Table S3**). In the DIABLO model, the metabolic dataset had the highest block weight (0.52) suggesting that it contributed most to classifying RS and NRS, while the lipidome, microbiome and PGS had similar weights (**Fig. 3 B**). This was supported by the sPLS-DA of the metabolic data resulting in the best performance measures (**Fig. 3 C**). The sPLS-DA (**Fig. 3 D**) confirmed the top features of the DIABLO model and pointed out further relevant microbes and lipid species that differed between RS and NRS (**Fig. 3 A**, **Table S1**).

**Figure 3:**
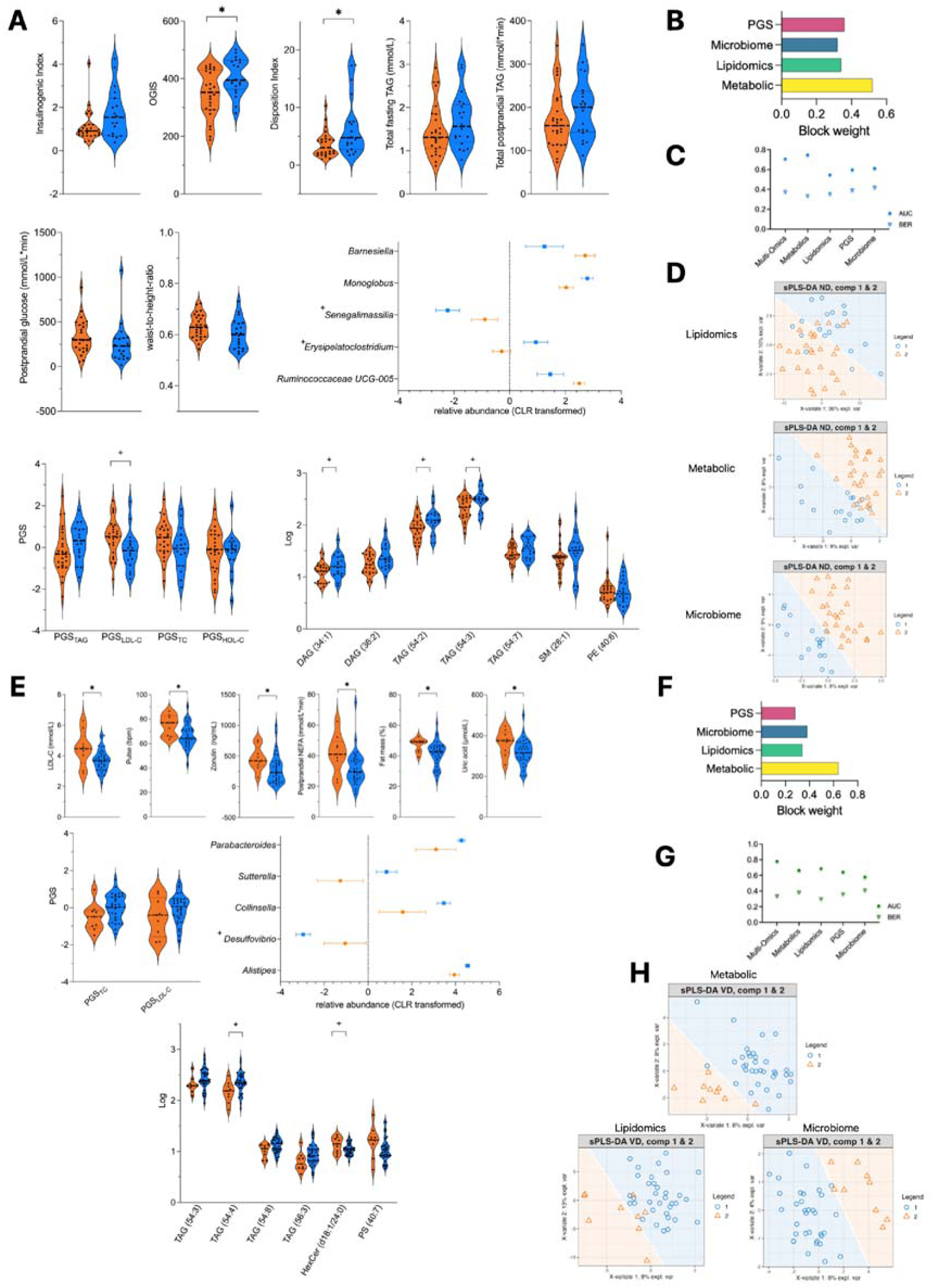
Models distinguishing RS and NRS in ND and VD at baseline. A: Violin and forest plots show levels or relative abundances + SEM (raw values for metabolic dataset, log transformed data for lipidomic dataset, CLR transformed data for microbiome dataset) of the top selected features from DIABLO, sPLS-DA and XGBoost models distinguishing RS (orange, *n*=27) and NRS (blue, *n*=20) from ND baseline. For visualization purposes, outliers of the insulinogenic and disposition index are not shown. Between-group differences were analyzed using Student’s t-test for metabolic data, linear regression or zero-inflated gaussian models for lipidomic data and Wilcoxon test for microbiome data. \**p*_adj_<0.05, +*p*_unadj_<0.05. B: Bar plot show contribution (block weight) of the four omics datasets to the component of the DIABLO model for ND. C: Plot shows the performance, measured by AUC and BER of the multi-omics (DIABLO) and single-omics (sPLS-DA) analyses distinguishing RS and NRS from ND at baseline. D: Sample plots from sPLS-DA include a prediction background to show the class assignment to RS (orange) or NRS (blue) for new data points based on their values on the first two latent components for the single-omics datasets. For metabolic, lipidomic and microbiome datasets, only components 1 and 2 are shown. No plot is shown for PGS, as it only has 1 component. E: Violin and forest plots show levels or relative abundances + SEM (raw values for metabolic dataset, log transformed data for lipidomic dataset, CLR transformed data for microbiome dataset) of the top selected features from DIABLO, sPLS-DA and XGBoost models distinguishing RS (orange, *n*=9) and NRS (blue, *n*=36) from VD combined at baseline. Between-group differences were analyzed using Student’s t-test for metabolic data, linear regression or zero-inflated gaussian models for lipidomic data and Wilcoxon test for microbiome data. \**p*_adj_<0.05, +*p*_unadj_<0.05. F: Bar plot show contribution (block weight) of the four omics datasets to the component of the DIABLO model for VD. G: Plot shows the performance, measured by AUC and BER of the multi-omics (DIABLO) and single-omics (sPLS-DA) analyses distinguishing RS and NRS from VD at baseline. H: Sample plots from sPLS-DA include a prediction background to show the class assignment to RS (orange) or NRS (blue) for new data points based on their values on the first two latent components for the single-omics datasets. For metabolic and microbiome datasets, only components 1 and 2 are shown. There is no plot for PGS as it only has 1 component. See also Table 1, S1, S2, S3, S4. *AUC, area under the curve; BER, balanced error rate; DAG, diacylglycerides; DIABLO, Data Integration Analysis for Biomarker discovery using Latent variable approaches for Omics studies; HDL-C, high-density lipoprotein cholesterol; HexCer, hexosylceramides; LDL-C, low-density lipoprotein cholesterol; LPC, lysophosphatidylcholines; ND, Nordic Diet; NEFA, non-esterified fatty acids; NRS, Non-Responders; OGIS, oral glucose insulin sensitivity index; p*_adj_, *adjusted p-value; PE, phosphatidylethanolamines; PGS, polygenetic risk scores; PS, phosphatidylserines; RS, Responders; p_unadj_, unadjusted p-value; SEM, standard error of mean; SM, sphingomyelins; sPLS-DA, sparse Partial Least Squares - Discriminant Analysis; TAG, triacylglycerides; TC, total cholesterol; VD, Vegetarian Diet*.

In confirmation of some of the most relevant features of the DIABLO model, the XGBoost multi-omics prediction model (sensitivity: 0.556, specificity: 0.741, F1-score: 0.571, AUC-ROC: 0.749) revealed PGS_LDL-C_, insulinogenic index, SM (28:1) and OGIS as the strongest predictors (**Fig. 3 A, Fig. S2 B, Table S4**). Of the single-omics datasets, the PGS led to the best prediction results, again confirming the PGS_LDL-C_ as the strongest predicting factor (**Table S4**).

These analyses point out that RS of the ND at baseline exhibit an inferior postprandial glucose metabolism as indicated by a lower insulin sensitivity and a higher genetic risk for hyperlipidemia compared to NRS.

### RS to the Vegetarian Diet exhibit an inferior clinical lipid profile, gut barrier integrity and body composition in comparison to NRS

For VD, *Alistipes*, LDL-C, pulse, zonulin, postprandial NEFA, fat mass, uric acid, TAG (54:4) and the PGS_TC_ and PGS_LDL-C_ were the strongest baseline features in the DIABLO model (AUC=0.779, *p*=0.028, BER_centroids_=0.333). Tailored univariate analyses confirmed higher levels of LDL-C (*p*_unadj_=0.012), pulse (*p*_unadj_=0.02), zonulin (*p*_unadj_=0.017), postprandial NEFA (*p*_unadj_=0.036), fat mass (*p*_unadj_=0.046) and uric acid (*p*_unadj_=0.046) in RS of the VD at baseline and lower levels of TAG (54:4) (*p*_unadj_=0.02) (**Fig. 3 E**, **Table S1, Table S2**). Univariate correlation analysis showed that fat mass correlated mildly with postprandial NEFA and PGS_TC_ and more strongly with zonulin levels. In the DIABLO model, the metabolic dataset contributed most to classifying RS and NRS with the highest block weight (0.64), followed by the microbiome and lipidome (**Fig. 3 F**, **Table S3**). In the sPLS-DA models, the best classification performance was achieved by the lipidomic dataset (**Fig. 3 G**). The single-omics analysis (**Fig. 3 H**) confirmed the top features from the multi-omics models as the most relevant factors and identified additional relevant microbes and lipid species (**Fig. 3 E**, **Table S1**).

Supporting the results of die DIABLO model, the XGBoost multi-omics prediction model for the VD (sensitivity: 0.912, specificity: 0.222, F1-score: 0.861, AUC-ROC: 0.541) identified pulse, fat mass, PGS_TC_ and LDL-C as well as phosphatidylserine (PS) (40:7) and TAG (54:3) as new features, as the strongest predictors (**Fig. 3 E, Fig. S2 C, Table S4**). Of the single-omics datasets, the lipidomics led to the best prediction results (**Table S4**).

These results indicate that RS of the VD are characterized by a less favorable profile in clinical markers of lipid metabolism, gut barrier integrity and body composition.

#### Multi-omics signature in diet-induced changes

To identify multi-omics signatures in the diet-induced changes that distinguish RS and NRS, we ran additional sPLS-DA and DIABLO models. In the combined group, changes in *Akkermansia, Lachnospiraceae NK4A136* group, LDL-C, total postprandial TAG, cholesteryl ester (CE) (20:3), TAG (56:8), TAG (56:6) and TAG (54:7) and the PGS_LDL-C_ and PGS_TC_ were the strongest features in the DIABLO model (AUC=0.747, *p*=0.011, BER_centroids_=0.130) (**Fig. 4 A-C**, **Table S1**). Tailored univariate analyses of the top features confirmed that besides the decrease in LDL-C (-17.7%, *p_un_*_adj_<0.001), RS were characterized by a reduction in CE (20:3) and an increase in TAG (56:8) (all *p*_unadj_≤0.006) (**Fig. 4 B, Table S5**). The change in LDL-C was strongly inversely correlated with the change in *Akkermansia* and with the PGS_LDL-C_ and PGS_TC_ (r≥-0.95) and positively with the change in CE (20:3) (r=1), which was mostly confirmed by Pearson correlation analysis. The change in total postprandial TAG was highly correlated with the change in all TAG species selected by the model, PGS_HDL-C_, and *Lachnospiraceae NK4A136* group (r≥0.7) (confirmed for TAG (56:6) by Pearson correlation analysis) (**Fig. 4 A**, **Table S3**). The metabolic dataset contributed most to the classification of RS and NRS, as shown by the block weight for component 1 (0.78) (**Fig. 4 D**) and the best classification metrics of the single-omics datasets (**Fig. 4 E)**. The sPLS-DA models (**Fig. 4 F**) confirmed the most important features of the DIABLO model and identified other microbes and lipid species that differentiate between RS and NRS (**Fig. 4 B**, **Table S1**).

**Figure 4:**
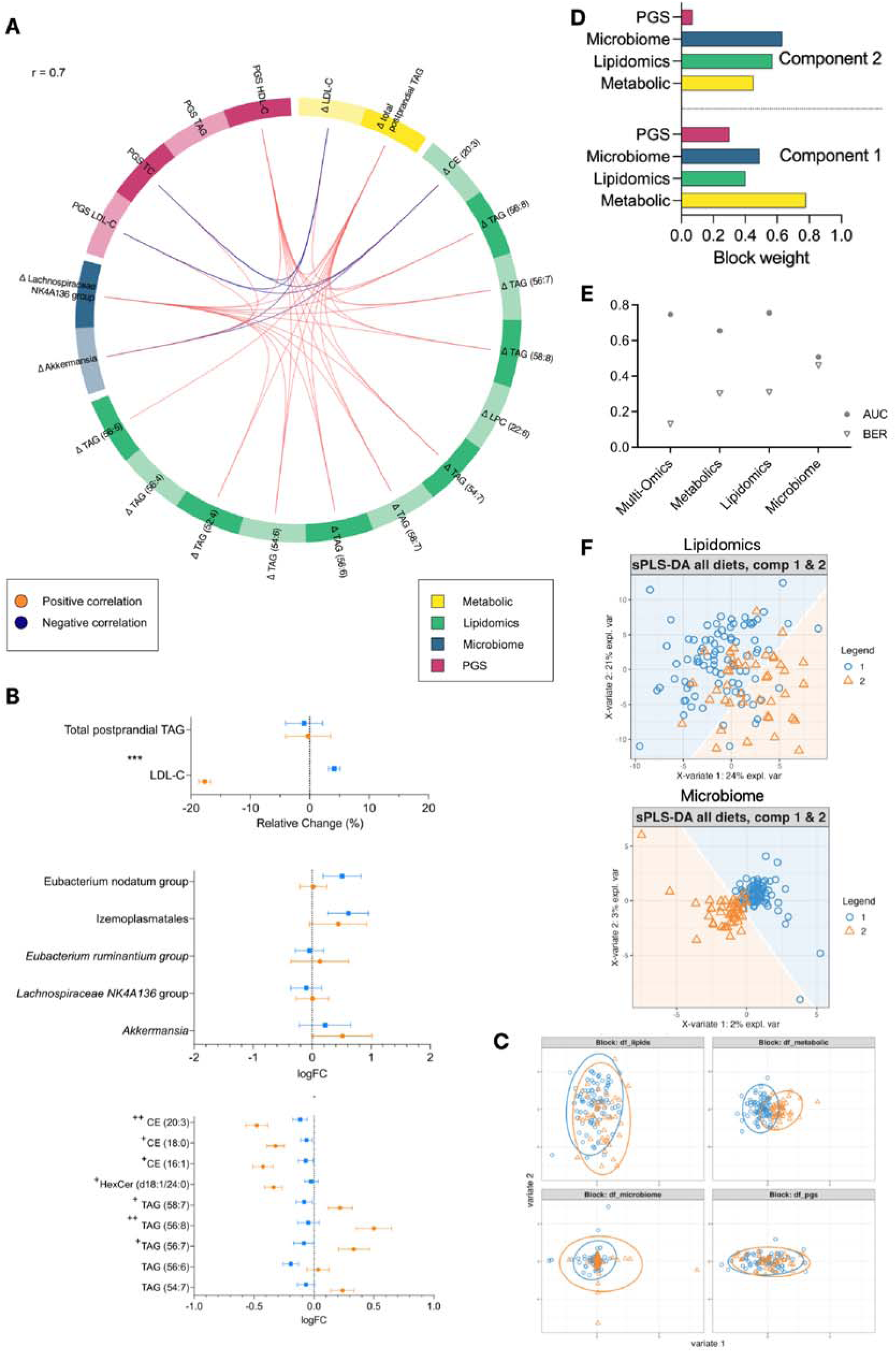
Models distinguishing the change in multi-omics features of RS and NRS in all diets combined. A: Circos plot shows positive (red) and negative (blue) correlations (cut-off: r=0.7) between the variables selected in the best DIABLO model characterizing the change in multi-omics features of RS and NRS from all diets combined. Four different omics datasets have been included: metabolic (yellow), lipidomic (green), PGS (blue) and gut microbiome (pink). B: Forest plots show relative change or log2fold change + SEM of the top selected features from DIABLO and sPLS-DA models distinguishing the change in multi-omics features of RS (orange, *n*=46) and NRS (blue, *n*=92) from all diets combined. Between-group differences were analyzed with linear mixed-models for metabolic data, linear regression or zero-inflated gaussian models for lipidomic data and Wilcoxon test for microbiome data. \**p*_adj_<0.05, \*\*\**p*_adj_<0.001. C: Sample plots show the samples projected onto the first two latent components obtained from the four integrated multi-omics datasets for RS (orange) and NRS (blue) in two components. D: Bar plot show contribution (block weight) of the four omics datasets to each component of the DIABLO model. E: Plot shows the performance, measured by AUC and BER of the multi-omics (DIABLO) and single-omics (sPLS-DA) analyses distinguishing the change in multi-omics features of RS and NRS from all diets combined. F: Sample plots from sPLS-DA include a prediction background to show the class assignment to RS (orange) or NRS (blue) for new data points based on their values on the first two latent components for the single-omics datasets. For microbiome and lipidomics only components 1 and 2 are shown. There is no plot for the metabolic dataset, because it only has 1 component. See also Table S1, S3, S5. *AUC, area under the curve; BER, balanced error rate; CE, cholesteryl esters; DIABLO, Data Integration Analysis for Biomarker discovery using Latent variable approaches for Omics studies; HDL-C, high-density lipoprotein cholesterol; LDL-C, low-density lipoprotein cholesterol; logFC, log fold change; NRS, Non-Responders; p*_adj_, *adjusted p-value; PGS, polygenetic risk scores; RS, Responders; SEM, standard error of mean; sPLS-DA, sparse Partial Least Squares - Discriminant Analysis; TAG, triacylglycerides; TC, total cholesterol*.

These results show that the LDL-C response in general is inversely associated with changes in *Akkermansia* and influenced by the polygenetic risk. Moreover, alterations in the lipidomic profile, specifically of long-chain TAG species are linked to the changes in the clinical lipid profile as well as specific gut microbes.

### LDL-C reduction in RS induced by the Nordic Diet is linked to changes in the lipidomic profile and gut microbiome

In the ND group, RS and NRS were classified by a DIABLO model with three components (AUC=0.812, *p*=0.005, BER_mahalanobis_=0.131). The strongest contributors to classification for component one were changes in *Oscillibacter*, LDL-C, SM (18:1), SM (24:2) and lyso-phosphatidylcholine (LPC) (20:4) and the PGS_TAG_. In component two, changes in *Bilophila, Megasphaera, Ruminococcus* gnavus group, postprandial NEFA, creatinine, in phosphatidylethanolamine (PE) (38:3), PE (38:4) and ether-linked phosphatidylethanolamine (PE-O) (40:7) and the age and the PGS_LDL-C_ and PGS_TC_ were the most important features. Component three included changes in *Butyricimonas*, *Megamonas* and *Eubacterium eligens* group, in acetic acid, fat mass, total postprandial TAG, CE (20:1), CE (20:4) and TAG (56:3) and the PGS_HDL-C_, PGS_TC_ and PGS_LDL-C_ as the most important features (**Fig. 5 A-C**, **Table S1**). Tailored univariate analysis showed that besides the stronger reduction in LDL-C (-18.9%, *p_un_*_adj_<0.001), RS also demonstrated a stronger reduction of HDL-C (-7.1%, *p_un_*_adj_=0.014) as well as an increase in SM (24:2) (*p*_unadj_=0.026) and *Eubacterium eligens* group (*p*_unadj_=0.042) (**Fig. 5 B, Table S5**). The change in LDL-C was strongly inversely correlated with the changes in SM lipid species (SM (18:1), (20:1), (24:1), (24:2), and with LPC (20:4) (all r≥-0.83) and positively with the change in TC, *Oscillibacter* and with PGS_TAG_ (all r≥0.93) (**Fig. 5 A**), which was largely confirmed by Pearson correlation analysis. The change in *Megasphaera* correlated negatively with the changes in the selected PE and PE-O lipid species (all r≥-0.58) and in *Parasutterella* (r=-0.87) and positively with the changes in *Ruminococcus* gnavus group, *Bilophila* and with the PGS_LDL-C_ and PGS_TC_ (all r≥0.82) (**Fig. 5 A**), which was broadly validated by Pearson correlation analysis (**Table S3**). In the DIABLO model the metabolic dataset had the highest block weight for components 1 and 3 (0.76 and 0.32) and the PGS for component 2 (0.36), suggesting that these datasets contributed most to classifying RS and NRS (**Fig. 5 D**). Of the single-omics data, the change in the gut microbiome discriminated RS and NRS of the ND best (**Fig. 5 E**). The sPLS-DA models (**Fig. 5 F**) confirmed most of the top features from the DIABLO model and identified additional relevant microbes (**Fig. 5 B**, **Table S1**).

**Figure 5:**
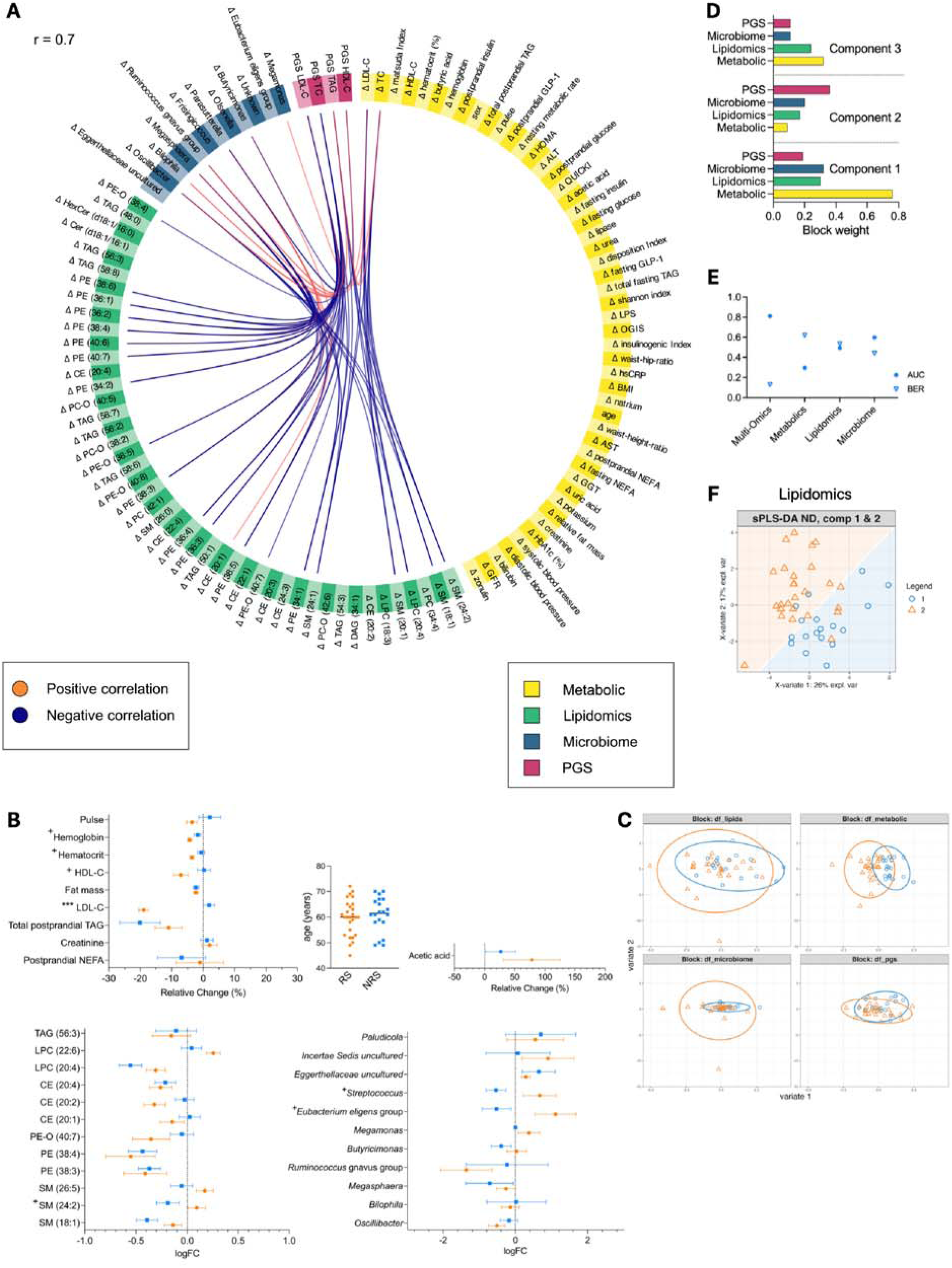
Models distinguishing the change in multi-omics features of RS and NRS in ND. A: Circos plot shows positive (red) and negative (blue) correlations (cut-off: r=0.7) between the variables selected in the best DIABLO model characterizing the change in multi-omics features of RS and NRS of ND. Four different omics datasets have been included: metabolic (yellow), lipidomic (green), PGS (blue) and gut microbiome (pink). B: Forest plots show relative change or log2fold change + SEM of the top selected features from DIABLO and sPLS-DA models distinguishing the change in multi-omics features of RS (orange, *n*=27) and NRS (blue, *n*=20) from ND. Interleaved scatter plot shows the age of RS and NRS. Between-group differences were analyzed with linear mixed-models for metabolic data, linear regression or zero-inflated gaussian models for lipidomic data and Wilcoxon test for microbiome data. \*\*\**p*_adj_<0.001, +*p*_unadj_<0.05. C: Sample plots show the samples projected onto the first two latent components obtained from the four integrated multi-omics datasets for RS (orange) and NRS (blue) in two components. D: Bar plot show contribution (block weight) of the four omics datasets to each component of the DIABLO model. E: Plot shows the performance, measured by AUC and BER of the multi-omics (DIABLO) and single-omics (sPLS-DA) analyses distinguishing the change in multi-omics features of RS and NRS from ND. F: Sample plot from sPLS-DA include a prediction background to show the class assignment to RS (orange) or NRS (blue) for new data points based on their values on the first two latent components for lipidomics (only components 1 and 2 are shown). There is no plot for the metabolic and microbiome datasets because they only have 1 component. See also Table S1, S3, S5. *ALT, alanine transaminase; AST, aspartate transaminase; AUC, area under the curve; BER, balanced error rate; CE, cholesteryl esters; Cer, ceramides; DIABLO, Data Integration Analysis for Biomarker discovery using Latent variable approaches for Omics studies; GFR, glomerular filtration rate; GGT, gamma-glutamyl transferase; GLP-1, glucagon-like peptide 1; HbA1c, hemoglobin A1c; HDL-C, high-density lipoprotein cholesterol; HexCer, hexosylceramides; HOMA, homeostatic model assessment; hsCRP, high sensitivity C-reactive protein; LDL-C, low-density lipoprotein cholesterol; logFC, log fold change; LPC, lysophosphatidylcholines; LPS, lipopolysaccharide; ND, Nordic Diet; NEFA, non-esterified fatty acids; NRS, Non-Responders; OGIS, oral glucose insulin sensitivity index; p*_adj_, *adjusted p-value; PC, phosphatidylcholines; PC-O, ether-linked phosphatidylcholines; PE, phosphatidylethanolamines; PE-O, ether-linked phosphatidylethanolamines; PGS, polygenetic risk scores; p*_unadj_, *unadjusted p-value; QUICKI, quantitative insulin sensitivity check index; RS, Responders; SEM, standard error of mean; SM, sphingomyelins; sPLS-DA, sparse Partial Least Squares - Discriminant Analysis; TAG, triacylglycerides; TC, total cholesterol*.

In conclusion, the LDL-C response to the ND is linked to changes in the gut microbiome and the lipidomic profile, specifically to *Oscillibacter* and SM lipid species, and is modified by the PGS.

### Vegetarian Diet-induced LDL-C reduction in RS is connected to the lipidomic profile and the gut microbiome

For VD, key features distinguishing RS and NRS were changes in *Coprobacter*, *Erysipelotrichaceae* uncultured, *CAG-56, Tyzzerella,* LDL-C, TC, fasting GLP-1, fasting NEFA, Potassium, Disposition Index, hexosylceramide (HexCer) (d18:1/24:0), HexCer (d18:1/16:0), LPC (20:1), LPC-O (16:0) and all four PGS in a DIABLO model (AUC=0.894, *p*=0.015, BER_mahalanobis_=0.181) (**Fig. 6 A-C**, **Table S1**). Tailored univariate analysis showed that besides the stronger reduction in LDL-C (-15.6%, *p_un_*_adj_<0.001), and TC (-9.3%, *p_un_*_adj_<0.001), RS also had a stronger reduction in NEFA (*p*_unadj_=0.047) and HexCer (d18:1/24:0) (*p*_unadj_=0.036) levels and an increase in fasting GLP-1 (*p*_unadj_=0.019) (**Fig. 6 B, Table S5**). In the DIABLO model, the change in LDL-C correlated positively with the change in all selected lipid species and especially strongly (all r≥0.82) with the change in HexCer (d18:1/24:0), TC, *Coprobacter* and with PGS_TC_ and PGS_LDL-C_ (**Fig. 6 A**). Pearson correlation analysis confirmed the correlation between LDL-C and TC with *Coprobacter* (**Table S3**). The metabolic and lipidomic dataset had the highest contribution (block weight) to component 1 (0.72) and component 2 (0.37), respectively (**Fig. 6 D**). In the sPLS-DA analyses, the performance of the metabolic, lipidomic and microbiome dataset were similar (**Fig. 6 E**, **Fig. 6 F**). Top features of the DIABLO model were confirmed and additional relevant microbes and lipid species were identified (**Fig. 6 B**, **Table S1**).

**Figure 6:**
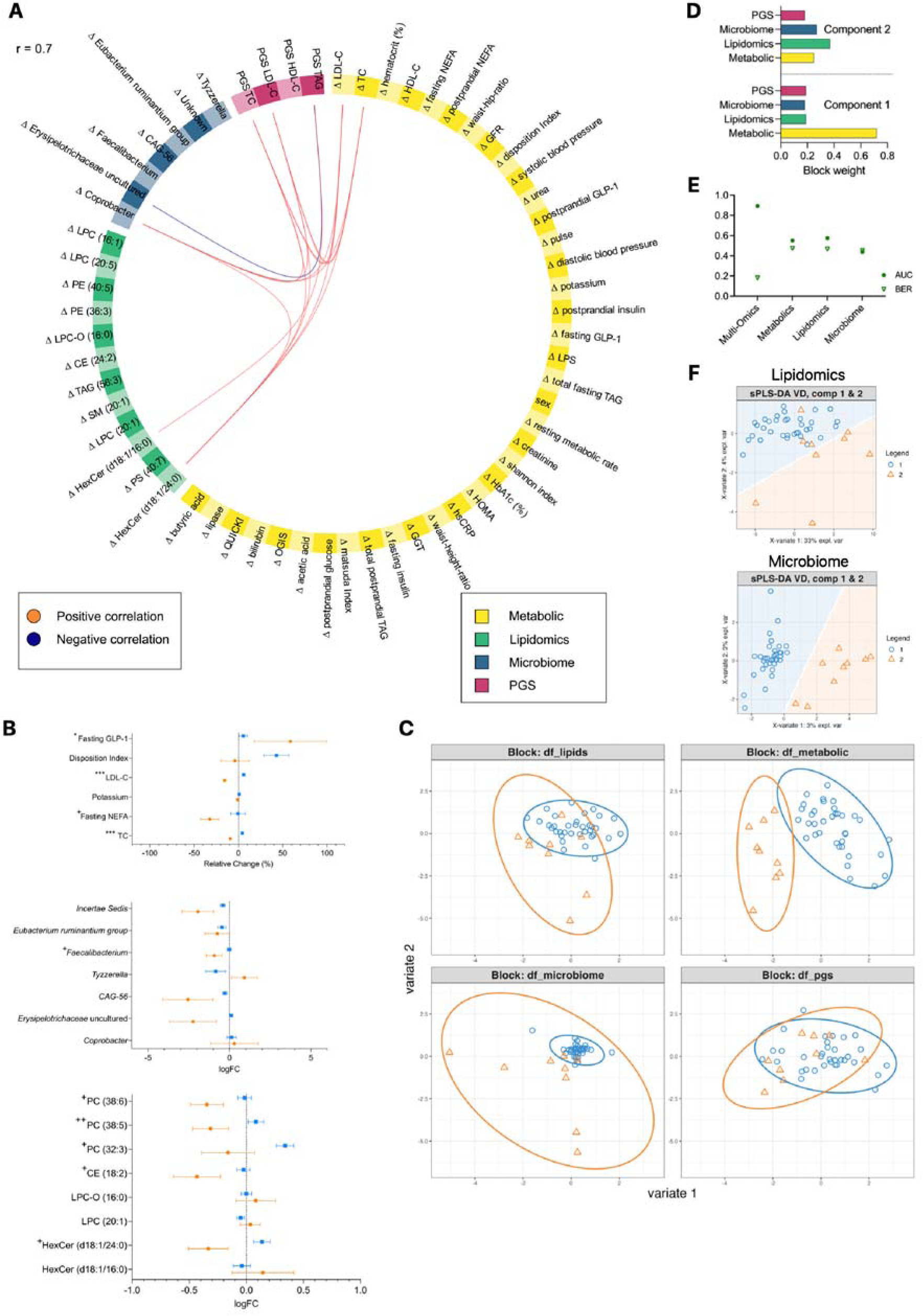
Models distinguishing the change in multi-omics features of RS and NRS in VD. A: Circos plot shows positive (red) and negative (blue) correlations (cut-off: r=0.7) between the variables selected in the best DIABLO model characterizing the change in multi-omics features of RS and NRS of VD. Four different omics datasets have been included: metabolic (yellow), lipidomic (green), PGS (blue) and gut microbiome (pink). B: Forest plots show relative change or log2fold change + SEM of the top selected features from DIABLO and sPLS-DA models distinguishing the change in multi-omics features of RS (orange, *n*=9) and NRS (blue, *n*=36) from VD. For visualization purposes, outliers of the disposition index are not shown. Between-group differences were analyzed with linear mixed-models for metabolic data, linear regression or zero-inflated gaussian models for lipidomic data and Wilcoxon test for microbiome data. \**p*_adj_<0.05, \*\*\**p*_adj_<0.001, +*p*_unadj_<0.05, ++*p*_unadj_<0.01. C: Sample plots show the samples projected onto the first two latent components obtained from the four integrated multi-omics datasets for RS (orange) and NRS (blue) in two components. D: Bar plot show contribution (block weight) of the four omics datasets to each component of the DIABLO model. E: Plot shows the performance, measured by AUC and BER of the multi-omics (DIABLO) and single-omics (sPLS-DA) analyses distinguishing the change in multi-omics features of RS and NRS from ND. F: Sample plot from sPLS-DA include a prediction background to show the class assignment to RS (orange) or NRS (blue) for new data points based on their values on the first two latent components for for lipidomics and microbiome (only components 1 and 2 are shown). There is no plot for the metabolic dataset as it has only 1 component. See also Table S1, S3, S5. *AUC, area under the curve; BER, balanced error rate; CE, cholesteryl esters; DIABLO, Data Integration Analysis for Biomarker discovery using Latent variable approaches for Omics studies; GFR, glomerular filtration rate; GGT, gamma-glutamyl transferase; GLP-1, glucagon-like peptide 1; HbA1c, hemoglobin A1c; HDL-C, high-density lipoprotein cholesterol; HexCer, hexosylceramides; HOMA, homeostatic model assessment; hsCRP, high sensitivity C-reactive protein; LDL-C, low-density lipoprotein cholesterol; logFC, log fold change; LPC, lysophosphatidylcholines; LPC-O, ether-linked lysophosphatidylcholines; LPS, lipopolysaccharide; ND, Nordic Diet; NEFA, non-esterified fatty acids; NRS, Non-Responders; OGIS, oral glucose insulin sensitivity index; p*_adj_, *adjusted p-value; PC, phosphatidylcholines; PE, phosphatidylethanolamines; PGS, polygenetic risk scores; p*_unadj_, *unadjusted p-value; PS, phosphatidylserines; QUICKI, quantitative insulin sensitivity check index; RS, Responders; SEM, standard error of mean; SM, sphingomyelins; sPLS-DA, sparse Partial Least Squares - Discriminant Analysis; TAG, triacylglycerides; TC, total cholesterol*.

In conclusion, the response to the VD is linked to changes in the lipidomic profile and gut microbiome, especially to HexCer (d18:1/24:0) and *Coprobacter* and to the genetic risk. Moreover, RS are characterized by an increase in GLP-1, which might be involved in the improvements in the clinical lipid profile^38^.

## DISCUSSION

In this study, we found marked individual differences in the LDL-C response upon two different dietary interventions based on established plant-based dietary patterns. In general, the ND led to the highest number of LDL-C RS. Of the four omics datasets, the metabolic and lipidomic profile at baseline in general classified RS and NRS best. There were specific features in the multi-omics profile at baseline and in the diet-induced changes that distinguish RS from NRS. RS irrespective of the dietary intervention were characterized by a less favorable initial clinical lipid profile, which became particularly visible in RS of the VD. RS of the ND demonstrated an inferior postprandial glucose metabolism at baseline, compared to NRS. Furthermore, the analysis of multi-omics data identified a particular pattern of features that classify RS and NRS, supporting our hypothesis that the interaction of specific features derived from four omics datasets may play a role. The detected patterns were characterized by a strong connection of metabolic, lipidomic and microbial features and the PGS.

Independent of the specific dietary pattern, several factors influencing the likeliness of reducing LDL-C levels through a dietary intervention were detected. Certain features such as higher baseline levels of TC and LDL-C, a lower abundance of *Parabacteroides,* specific lipid species, especially TAG, as well as a lower insulinogenic index, were reflected across multiple models and therefore seem to play an important role for the characterization of RS.

Our results indicate that persons with a less favorable clinical lipid profile are more likely to respond to a dietary intervention, which is especially relevant as increased lipid levels are strongly linked to an elevated risk for atherosclerosis and cardiovascular diseases^19,21,39–41^ and these persons can benefit most from a dietary intervention resulting in improved lipid levels, as this is directly associated with a significant disease risk reduction^23,42–44^. Our finding that individuals in the combined group and VD with higher initial LDL-C or TC values profited more from an intervention is supported by results of the Food4Me study^10^ and was also observed in children with hyperlipidemia after a 6-months Mediterranean diet^45^. In our study, the baseline levels of LDL-C were linked to the PGS for hyperlipidemia, which are highly associated with CVD risk. Therefore, in persons with higher PGS, interventions aiming to reduce lipid levels are especially relevant to compensate for the genetic predisposition^46,47^, as adopting a healthy lifestyle can mitigate the elevated risk by up to 50%^47^ and might prevent clinical disease manifestations and the need for cholesterol lowering medication^48^. Our results show that especially persons with higher PGS for LDL-C profit from a ND intervention, underlining our previous results^49^.

TAG and DAG lipids are involved in various physiological processes, including intracellular insulin signaling, secretion and in the development of insulin resistance^50^. Total levels of TAG and DAG are generally higher in individuals with dyslipidemia and metabolic syndrome (MetS), and associated with glucose levels^51^. Thus, in the ND, total fasting and postprandial TAG were among the top features to distinguish RS and NRS. Despite having comparable (or only slightly lower) levels of total fasting and postprandial TAG compared to NRS, RS of the ND and VD group had lower levels of TAG lipid species (54:2), (54:3), (54:4) and DAG (34:1), demonstrating that RS are characterized by a specific lipidomic profile. These results point out the importance of assessing the lipid profile in-depth, since by only focusing on the total amount of TAG, differences that are present on a deeper level might be overlooked. Moreover, lipid species have different functions and effects in the body and may show varying associations with diseases^16,51–53^. Additionally, our results indicate that features such as total TAG are of relevance in connection with other omics-features, even when mean values are similar between RS and NRS. In accordance with this, we detected specific gut microbes relevant for the characteristic multi-omics signature, while the overall gut microbiome diversity and composition between RS and NRS were comparable. In the ND group, total TAG levels correlated positively with specific lipid species such as TAG (54:5), DAG (36:2), SM (28:1) and PE (40:6) and inversely with *Ruminococcaceae UCG-005,* suggesting a possible link between the gut microbiome and lipid metabolism. In line with this, *Ruminococcaceae UCG-005* has been associated with lipid metabolism and obesity-related parameters in rats fed a high-fat diet. There the diet-induced increase in lipid levels of TAG, TC and LDL-C correlated with a depletion of *Ruminococcaceae UCG-005,* indicating that the dietary effect was in part linked to changes in the microbiome^54^, as also shown in our results.

Furthermore, there were diet-group specific features characterizing RS in the VD like higher levels of fat mass, zonulin, uric acid, pulse and postprandial NEFA. The top features in the VD point out the central role of obesity, linking impairments in gut barrier integrity^55^ with metabolic alterations, in particular lipid metabolism^56,57^ and uric acid^58^, and with pulse as a marker of cardiometabolic health^59^. Obesity is a driver of insulin resistance and hyperlipidemia, mainly through adipose tissue dysfunction and chronic low-grade inflammation^60^, contributing to further metabolic dysfunction, hypertension and the development of secondary diseases^61,62^. Therefore, improving the lipid profile via personalized dietary interventions in persons with an obesogenic profile is especially relevant to counter metabolic disturbances and prevent further diseases.

Additionally, the known close link between lipid and glucose metabolism^63–70^ was also evident in our study, where RS of the ND and in the combined group showed poorer insulin sensitivity compared to NRS, which has been associated with an atherogenic profile characterized by elevated total TAG and NEFA levels, reduced HDL-C levels, a shift in LDL-C composition, and with the lipidomic profile^63,71^. Our results and those of other intervention studies, showing that tissue-specific insulin resistance phenotypes influence the effect of specific dietary interventions, underline the importance of the initial insulin sensitivity on the cardiometabolic response to targeted dietary interventions^72,73^. Several underlying mechanisms of insulin resistance, such as the activation of protein kinase C θ (PKCθ), lead to a disruption of the normal insulin signaling cascade and thus an impairment in glucose uptake and utilization by muscle cells involving specific lipid species like DAG and ceramides (CER)^71,74^. Therefore, the lipid species, e.g., DAG (34:1) and (36:2), that were discriminative of RS of the ND likely play a role linking the less-favorable initial insulin sensitivity with greater improvements in lipid metabolism. Besides, the correlation of *Ruminococcaceae UCG-005* with the measures of insulin sensitivity support the relevance of this genus in determining the response in LDL-C in the ND and the mediating role of the gut microbiome.

Our results indicate that the initial state of the lipid metabolism, the gut barrier integrity and the body composition are highly relevant for responding to the VD, while the initial postprandial glucose metabolism, specifically insulin sensitivity, is important for the response to the ND. Specific gut microbes and lipid species play a key role linking diet and metabolism. These diet-specific features may help identify persons which will likely profit from changing their habitual diet to a tailored diet such as VD or ND. Even though some of the specific characteristics differ between RS of the VD and the ND, they generally represent a less favorable cardiometabolic profile of RS compared to NRS, pointing out that dietary interventions are especially beneficial in persons with an elevated risk for the development of CVD. Improving the lipid profile through diet contributes to a better insulin sensitivity, disrupting the cycle of metabolic dysfunction, and might delay or prevent the onset of further impairments such as the progression to diabetes, and maintain health^61,71^. Moreover, our results emphasize that high-resolution multi-omics analyses are relevant for a comprehensive characterization and potential stratification of participants for targeted interventions in the context of personalized nutrition^7,75–78^. Strong individual variations might mask differences between groups, and especially the microbiome and lipidome are complex systems that we are just beginning to understand^51,79^. This complicates the assessment of their role in the dietary effects and requires adequate statistical tools.

In addition to the models assessing differences between RS and NRS at baseline, we performed further analyses investigating how diet-induced changes in multi-omics features are connected to the reduction in LDL-C that characterizes RS. Irrespective of the diet, an increase in *Akkermansia* abundance after intervention was one of the top features characterizing RS and correlated positively with the PGS_TC_, PGS_LDL-C_ and inversely with the reduction in LDL-C and CE (20:3), indicating that the genetic risk impacts the metabolic reaction to the intervention in interaction with the microbiome. The genus *Akkermansia* has been linked to improvements in metabolic health and glucose metabolism. Specifically, for the species *Akkermansia muciniphila (A. muciniphila)*, which was the most common species of this genus in our data, effects on glucose and lipid metabolism are well-established, and it is currently being investigated as a next-generation probiotic^80^. In support of our results, *A. muciniphila* supplementations led to reduced levels of TC in animal studies^81^ and to improvements in insulin sensitivity, insulinemia, TC levels, bodyweight and fat mass in humans^82^. Despite stable indices of insulin resistance in our RS, the substantial improvements in cholesterol levels might have contributed to an enhancement of insulin sensitivity over a longer intervention duration. In addition, RS of all diets had a stronger decrease in some CE lipid species, e.g., CE (16:1), (18:0) and (20:3). Higher levels of CE (20:3) have been measured in patients with diabetes^83^ as well as in mice fed a high-fat diet^84^, and have been associated with higher LDL-C and TAG values^41^. Conversely, a diet-induced reduction in CE (20:3) is beneficial^52,85^ and in line with our findings, supporting the importance of in-depth analysis. In addition, RS exhibited an increase in TAG species with higher acyl chain lengths and number of double bonds like TAG (56:7), (56:8) and (58:7), which have been linked to a lower CVD risk, while the opposite was shown for shorter TAG with fewer double bonds^52,86^. These diet-induced changes of TAG lipid species are of relevance since the number of carbon atoms and the number of double bonds are important for their biological function^51,87^. Overall, the changes in the lipidomic profile of RS reflect an improvement of CVD risk, relevant in the context of personalized nutrition.

In the ND, a decrease of *Oscillibacter* was among the strongest features describing RS, correlating highly with the reduction in LDL-C and inversely with the SM lipids. *Oscillibacter* has recently been identified as relevant in the context of cholesterol metabolism, since *Oscillibacter* is linked to lower levels of cholesterol in participants from the Framingham Heart Study^88^. Additionally, cholesterol-metabolizing capabilities of *Oscillibacter* gut isolates were demonstrated *in vitro*^88^. Thus, the reduction of 30% of *Oscillibacter* we found in our RS might be due to the diet-induced reduction of TC and LDL-C of 14.8% and 18.9% as metabolizable substrate. This hypothesis is supported by a study with hamsters, showing that a high-fat and high-cholesterol diet induced rising levels of TC and LDL-C as well as higher abundances of *Oscillibacter* while a supplementation with Oryzanol, a mix of ferulic acid esters of plant and phytosterols, reversed these changes^89^. Additionally, in an RCT, following a high-dose oat diet for two days led to substantial improvements in cholesterol levels in participants with MetS which were linked to phenolic compounds from oats, e.g., ferulic acid, as well as their microbial derivates^90^, suggesting a mechanistic connection. Since oats and berries, which are high in phenolic compounds^91,92^, were among the key foods of the ND, the reduction in *Oscillibacter,* LDL-C and TC we observed in ND RS may have been linked to these food items. In addition, the detected changes in the selected SM lipids inversely correlating with the reduction in LDL-C and *Oscillibacter* hint to another mechanistic link. Since SM lipids are central components of plasma lipoproteins and influence both their structure and function, they are involved in cholesterol absorption, transport and hepatic cholesterol metabolism. Moreover, a dietary supplementation of SM lipids has been shown to help retain homeostasis and may benefit the prevention of metabolic diseases^86,93^

Our results show that the 6-weeks ND and VD interventions improved clinical lipids and the lipidomic profile, in particular TAG, SM and CE lipids, in an individual manner, with the gut microbiome and the genetic risk for hyperlipidemia strongly impacting the individual LDL-C response. Specifically, *Akkermansia* and *Oscillibacter* were relevant genera that may have contributed to the reduction in LDL-C. Given the high interactions between the different features, especially a strong inter-relation between changes in metabolism with gut microbes and the lipidomic profile, multi-omics approaches are a promising strategy for the characterization of RS to dietary interventions.

Supporting the diet-dependent connection we found between the changes in lipid metabolism with the gut microbiome, different dietary fats have been shown to influence host-microbe interactions in metabolism and immune responses distinctly, mainly mediated by bile acids^94,95^. Both the ND as well as the VD are rich in mono-or polyunsaturated fatty acids as well as fiber and phytosterols^96,97^, which have been shown to significantly alter blood lipids, when replacing dietary saturated fatty acids^16,98–101^. Particularly omega-3 fatty acids, present in fatty fish, have a beneficial impact on lipid and glucose metabolism and prevent or reverse low-grade inflammation in adipose tissue and insulin resistance^71,102,103^, possibly explaining the higher number of RS we had in the ND compared to the VD. We assessed characteristics of RS to different dietary interventions that are based on health-promoting plant-based dietary patterns^34,35^. In addition to the metabolic improvements they induce^30^, they are increasingly relevant due to their advantages regarding environmental sustainability and in the context of climate change^104–106^. Investigating dietary patterns has certain advantages over assessing the effects of single meals, foods, or nutrients. For example, it provides the opportunity of considering combined and synergistic effects of dietary compounds, has more real-world applicability and stronger disease association^107,108^. Besides, the ND and VD patterns we investigated, are feasible, easy-to-implement and culturally acceptable diets^106,109–111^. However, not all participants profited equally from adopting these diets and we detected specific features at baseline that predicted the response. In support of our approach of assessing the multi-omics profile of RS and NRS, results of the Food4Me study suggest that the identification of phenotypic factors, influencing the response to a dietary intervention, is important for the advancement of precision nutrition^10^. Thus, knowing the likeliness of profiting from an intervention from the beginning might assist in adhering to the diet and in providing personalized dietary recommendations.

The analyses are based on a well-conducted randomized, controlled dietary intervention trial comparing the effects of three dietary patterns in a deeply phenotyped cohort of persons at elevated CVD risk. We applied single-as well as multi-omics approaches and combined prediction models with descriptive models and tailored univariate testing to investigate characteristics of RS and NRS in detail. In order to find the best performing model, we tested different design settings for all models and chose the design providing the lowest error rate. XGBoost, a state-of-the-art method and powerful machine learning tool was used for the prediction models^37,112^. We had more NRS than RS (except for the ND), resulting in an unbalanced dataset which we accounted for by reporting the BER^113^ for the spls-DA and DIABLO models and by including a parameter weighing RS and NRS in the XGBoost models. Moreover, our sample size of 138 was high for a nutritional intervention study, enabling us to detect signatures of specific multi-omics features that characterize RS and NRS to a ND and VD intervention, two plant-based and culturally acceptable dietary patterns^106,109–111^.

Our results show that RS and NRS to a dietary pattern intervention differ in their baseline multi-omics profile as well as in the diet-group specific changes after intervention. We identified key determinants influencing the individual differences in the dietary effects on LDL-C reduction, which is a central strategy in cardiovascular risk management. Moreover, we predicted the response to the interventions based on the baseline multi-omics profile. Persons with a less favorable clinical lipid, glucose or lipidomic profile, a higher polygenetic risk for hyperlipidemia and who carry specific bacteria at baseline, are more likely to profit from a dietary intervention than those with a healthier initial state, which is relevant in the context of their increased cardiometabolic risk. The plant-based dietary patterns we examined provide a realistic, beneficial and sustainable approach for a dietary intervention in a high-risk cohort. Moreover, we detected distinct patterns of features in the diet-induced changes. The beneficial effects on lipid metabolism of our dietary interventions were influenced by the genetic risk for hyperlipidemia and associated with changes in gut microbiome. These specific signatures we observed may be used as biomarkers for personalized nutrition approaches to improve the impact of dietary interventions on cardiovascular risk and to maintain health.

## RESOURCE AVAILABILITY

### Lead contact

Requests for further information and resources should be directed to and will be fulfilled by the lead contact, Alina Schieren.

### Materials availability

This study did not generate new unique reagents.

### Data and code availability

- Anonymized source data, including the raw lipidomic, microbiome 16S rRNA, PGS and metabolic data that support the findings of this study are provided with this paper or have been deposited on the Zenodo platform (https://zenodo.org/records/17535871). Access can be obtained from the corresponding author upon reasonable request.
- Publicly available R packages have been used for analyses in this study: mixOmics^114^, qiime2R and phyloseq. All original code has been deposited on the Zenodo platform (https://zenodo.org/records/17535871). Access can be obtained from the corresponding author upon request.
- Any additional information required to reanalyze the data reported in this paper is available from the lead contact upon request.

## Supporting information

Supplemental Items

## Data Availability

All data produced in the present study are available upon reasonable request to the authors

## ACKNOWLEDGEMENTS

The authors would like to thank the study participants for their participation in the trial, the lab technicians of the Departments Nutrition Physiology, Human Nutrition and Nutrition and Microbiota, University Bonn and the medical staff of the Central Laboratory, University Hospital Bonn for their great support in lab analyses.

The authors gratefully acknowledge the access to the Marvin high-performance computing (HPC) cluster of the University of Bonn and the Xcat high-performance computing (HPC) cluster of the University Hospital Bonn, Institute for Genomic Statistics and Bioinformatics & Institute for Medical Biometry, Informatics and Epidemiology, which was used for computationally demanding model fitting.

## AUTHOR CONTRIBUTIONS

A.S. and H.H. conducted the clinical trial. A.S., H.H., W.S., R.D.P, B. H., J.J.H. and M.Y. performed the laboratory analysis; A.S., C.A.G., and A.M. performed the statistical analysis. M.C.S., P.S., M.C., M.S., C.T., M.N. and J.H. supervised the trial and/or laboratory and data analysis. A.S. prepared the first draft of the manuscript, which was subsequently finalized in close collaboration with H.H., C.A.G., A.M., M.C.S. All authors provided substantial content contributions and edited the manuscript. A.S., C.A.G., A.M. and M.C.S. created and edited the Figures. All authors have read and approved the final manuscript.

Figure 1 A was created using Biorender. The icons displayed in Supplementary Figure S3 were made by kornkun, Those Icons, Eucalyp, Freepik, manshagraphics, Good Ware, SBTS2018, rizal2109 and DinosoftLabs from www.flaticon.com.

## DECLARATION OF GENERATIVE AI AND ASSISTED TECHNOLOGIES IN THE WRITING PROCESS

During the preparation of this work the authors used ChatGPT in order to check the grammar and language of parts of the manuscript and to improve the code. After using this tool, the authors reviewed and edited the content as needed and take full responsibility for the content of the published article.

## ADDITIONAL INFORMATION

Prior Presentation

Part of this research has been presented at the Precision Nutrition Forum and PREDIMED Omics Symposium 2025, Harvard University, Boston.

## INCLUSION AND DIVERSITY

We support inclusive, diverse, and equitable conduct of research.

## STAR METHODS

### Experimental Model and Study Participant Details

#### Study design

In this randomized, controlled intervention trial, two study phases of 6-weeks duration each, were conducted in parallel design. In the first study phase, conducted between November 2018 and April 2020, participants were randomly allocated to one of three groups (one control and two intervention groups): Habitual (western-type) control Diet (HD), lacto-ovo Vegetarian Diet (VD) or Nordic Diet (ND). Randomization tables in block format were generated by a statistician not involved in the clinical conduction of the study. After completion of the first study phase, participants were classified into Responders (RS) or Non-Responders (NRS) according to their change in LDL-cholesterol (LDL-C). NRS of the first study phase were subsequently invited to participate in the second study phase and were screened for eligibility again.

In the second study phase, which was conducted between October 2020 and December 2021, the participants were randomly allocated to one of the other two diet groups in a cross-over design. After completion of the second study phase, participants were once again classified into RS or NRS. Study visits were conducted at the study center of the Department of Nutrition and Microbiota, Institute of Nutritional and Food Sciences, University of Bonn, Germany, before and after intervention. On the study visits, anthropometric measurements, resting energy expenditure (indirect calorimetry), blood pressure and heart rate, deep metabolic characterization with assessment of clinical, metabolic, genetic and lipidomic parameters from fasting and postprandial blood samples obtained from an oral glucose tolerance test (OGTT), and fecal samples for analysis of the gut microbiome and short chain fatty acids were collected.

The study protocol was conducted according to the Declaration of Helsinki, approved by the ethics committee of the Medical Faculty of the University of Bonn, Germany (code 278/18) and the trial was prospectively registered at the German Clinical Trial Register (http://www.drks.de) under the identifier DRKS00015861. All participants provided written informed consent.

#### Participants

Details of the participants recruitment and randomization have been described previously^49^. Briefly, women and men of caucasian origin with overweight or obesity (BMI: 27-39.9 kg/m^2^), aged 45 to 70 years, were included into the study. Main inclusion criteria was the presence of at least one characteristic of the MetS as defined by (i) visceral fat distribution (waist circumference ≥80 cm for women, ≥94 cm for men), (ii) prehypertension (systolic blood pressure: ≥120-139 mmHg and/or diastolic blood pressure: ≥80-89 mmHg) or hypertension stage 1 (systolic blood pressure: ≥140-159 mmHg and/or diastolic blood pressure: ≥90-99 mmHg), (iii) dyslipidemia (fasting serum TAG ≥1.7 mmol/L and/or serum HDL-cholesterol <1.3 mmol/L for women and <1.0 mmol/L for men) or (iv) pro-inflammatory state (hs-CRP ≥2.0 mg/L). Among the reasons for exclusion were smoking, chronic diseases, type 2 diabetes, acute infections, chronic intake of dietary supplements, antibiotic treatment within four weeks before study participation, practiced vegetarianism or veganism or fish allergy, recent surgery or weight loss. Eligibility for study participation was checked within a screening visit comprising physical and clinical assessments as well as questionnaires. A flow diagram providing an overview of the progress of participants through the trial and the sample size can be found in **Fig. S1**.

#### Dietary interventions

Participants who were randomized into one of the intervention groups (VD or ND) received iso-energetic meal plans adapted to their caloric requirements (assessed by indirect calorimetry). Meal plans comprised characteristic food items of a ND or VD and consisted of three main meals and two snacks per day. Participants in the control group (HD) were instructed with adhering to their habitual (western-type) diet, without further specific guidelines regarding energy intake and food selection. All participants were advised to maintain their regular physical activity levels throughout study participation. Key components of a ND are foods traditionally consumed and available in Scandinavian countries such as root vegetables, potatoes, berries, wholegrain oats and rye, low-fat dairy products, rapeseed oil, nuts and fatty sea fish. The vegetarian study diet excluded any meat or fish products and consisted mainly of vegetables, legumes, fruits, wholegrain wheat products, high-fat milk, olive oil, seeds. A western-type diet comprises low intake of vegetables and fruit and high intake of processed foods, red meat, food rich in saturated fat and/or sugar and of sugar-sweetened beverages. Further details on the study diets have been described previously^49^.

### OGTT

An OGTT with a 3-hour observation period was performed after an overnight fast at baseline and after six weeks of intervention according to standard World Health Organization (WHO) guidelines^115^ with 75 g of glucose and 300 mL of water (Accu-Chek Dextrose O.G.T, Roche Diagnostics, GmbH, Mannheim, Germany).

#### Compliance

To assess compliance, participants from the intervention groups continuously documented their consumed meals and completed short-form questionnaires assessing the frequency of the consumption of the respective key food groups before and during study participation. Participants from the control group filled out two open 3-day dietary protocols (within week three and week six) to confirm their western-type dietary behavior throughout the study. Moreover, weekly phone calls with all study participants took place in addition to the regular study visits, in order to support with possible problems or questions and to increase compliance. Further, anthropometrics (monitoring of body weight, body composition and resting energy expenditure) (**Table S6**) and a variety of blood biomarkers (plasma concentrations of vitamins and phospholipid fatty acid composition) were assessed and confirmed compliance in the three diet groups as objective markers^116^.

#### Definition of Responders and Non-Responders

LDL-Cholesterol (LDL-C) was chosen as the decisive parameter for dividing participants into RS and NRS due to its strong association with the risk of CVD-related events^23,25,117^ and due to high inter-individual variations in the change of LDL-C. We conducted a literature search on the effects of Nordic and Vegetarian diets on LDL-C in order to set a cut-off value for the distinction of RS and NRS. Nordic Diets led to a reduction of 4-21%^31,118–121^ and Vegetarian Diets of around 10%,^122,123^ while most of them had longer intervention durations and reductions in body weight, which was kept stable in our study. Cholesterol lowering drugs can lead to reductions in LDL-C of 5-60% depending of the type and dose of medication^124^. Considering these effects observed in other nutritional studies and the fact that even modest reductions in LDL-C are considered clinically relevant^25,125^, we set a reduction of 10% as the cut-off value for our definition of RS. Accordingly, all participants with a reduction of 10% or more were defined as RS and all participants with a reduction of less than 10%, no change or an increase in LDL-C as NRS. A reduction of LDL-C of 10% in our study is equivalent to-0.38 mmol/L, about one third of the mean effect of statin therapy after one year of intake^25^. The word responder is used for participants of all three diet groups, but in the HD-group we understand this word as study RS, not diet RS.

## Method Details

### Measurements

A detailed description of the methods applied to measure anthropometrics (BMI, waist-height-ratio, waist-hip-ratio, fat mass), resting energy expenditure, blood pressure and heart rate, laboratory analyses of compliance, metabolic, lipidomic, genetic, fecal short chain fatty acids and microbiome data has been reported^49^. The next chapters comprise a brief description. Fasting and postprandial plasma and serum samples during a 3-h OGTT (after 15, 30, 45, 60, 120 and 180 minutes) and fecal samples were collected at baseline and after six weeks of intervention.

#### Metabolic data

The metabolic dataset comprises data from routine laboratory analyses measured in a certified laboratory (Central Laboratory of the Institute of Clinical Chemistry and Clinical Pharmacology at the University Hospital Bonn, Germany): fasting and postprandial glucose, insulin and TAG levels, fasting TC, LDL-C and HDL-C, levels of high sensitivity C-reactive protein (hsCRP) and hemoglobin A1c (HbA1c), clinical biochemistry (natrium, potassium, creatinine, glomerular filtration rate (GFR), urea, uric acid, bilirubin, gamma-glutamyl transferase (GGT), alanine transaminase (ALT), aspartate transaminase (AST), Lipase) and hematology (hemoglobin, hematocrit) parameters. Methods specifications are available online (https://www.ukbonn.de/ikckp/zentrallabor/leistungsverzeichnis/). Concentrations of non-esterified fatty acids (NEFA) from fasting and postprandial serum samples were measured using an in-vitro enzymatic colorimetric method assay (NEFA-HR (2), Wako Diagnostics, Mountain View, CA, USA) according to the manufacturer’s instructions (inter-assay/intra-assay variability 5.4%/4.8%) at the Institute of Nutrition and Food Sciences, University Bonn, Germany, as previously described^49^. Fasting and postprandial plasma concentrations of GLP-1 were analyzed with a total GLP-1 NL-ELISA (10-1278-01, Mercodia, Uppsala, Sweden) at the Department of Biomedical Sciences, University of Copenhagen, Denmark according to manufacturer’s instructions^126^. Lipopolysaccharide (LPS) and zonulin were measured in serum samples with ELISA (LPS: Cusabio Technology LLC, Houston, USA; zonulin: Antibodies-online GmbH, Aachen, Germany). Homeostasis Model Assessment (HOMA), Oral Glucose Insulin Sensitivity (OGIS), quantitative insulin sensitivity check index (QUICKI), Matsuda, Insulinogenic and Disposition index were calculated as measures of insulin resistance with the following formulas:

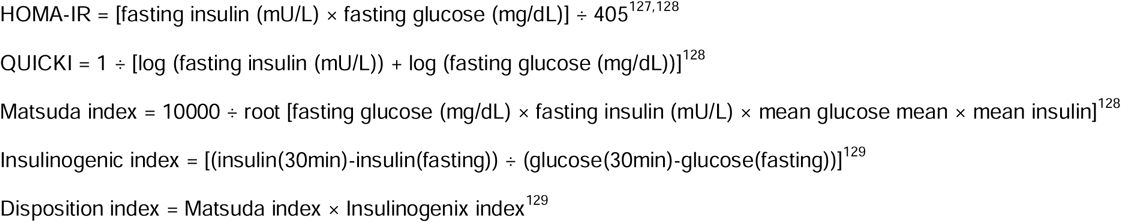

OGIS was calculated with a spreadsheet from Mari *et al.*^130^.

#### Lipidomics

Lipids from plasma samples were extracted by a modified Bligh & Dyer method. Briefly, 250 μL methanol:chloroform 1/1 containing 250 pmol PE 31:1(d7), 472 pmol PC 33:1(d7), 98 pmol PS 31:1, 56 pmol PA 31:1, 51 pmol PG 28:0, 39 pmol LPA 17:0, 35 pmol LPC 17:1, 38 pmol LPE 17:1, 32 pmol Cer 17:0, 241 pmol SM 18:1(d9), 55 pmol GlcCer 12:0, 339.7 pmol TG 50:1-D4, 740 pmol CE 18:1(d6), 64 pmol DG 31:1 and 103 pmol MG 17:1 as internal standard, was quickly added to one sample and followed by sonication for 1 minute in the bath sonicator. Subsequently, the samples were centrifuged at 20.000 g for 2 min and the supernatants were transferred into new tubes for further process. 300 µl chloroform and 700 µl 1% acetic acid in water were added into the samples to induce phase separation. The samples were shaken manually for 5 seconds and centrifuged at 20.000 g for 2 min. The upper phase was carefully removed and discarded. The entire lower phase was transferred into a new tube and evaporated in the centrifuge at 45 °C for 20 min. 1 mL spray buffer (2-propanol/methanol/water 8/5/1+10 mM ammonium acetate) was added into the samples and sonicated for 5 min.

Mass spectra was acquired using a Thermo Q-Exactive Plus spectrometer equipped with a standard heated ESI source. Direct infusion was performed with a Hamilton syringe driven by a syringe pump and managed via the Tune instrument control software. MS1 spectra with resolution 280,000 were recorded in 100 m/z windows from 250 – 1,200 m/z in positive mode followed by recording MS/MS spectra with resolution 70,000 by data independent acquisition in 1 m/z windows from 250 – 1,200 m/z in positive mode. The raw data were converted to.mzML files using MSConvert and analyzed with LipidXplorer software. For further analysis absolute amounts were calculated using internal standard intensities.

#### Microbiome analysis and short chain fatty acids

Fecal samples were collected at home one day before the baseline and endline visits and were brought to the study center. Upon arrival, a part of each sample was separated for the analysis of short chain fatty acids (butyric acid, pyruvic acid, lactic acid and acetic acid), as previously described^49^. The remaining samples were immediately stored at-80°C until further analysis. Pyruvic acid and lactic acid were excluded from the analysis because more than 50% of the values were below the detection limit. DNA extraction and high-throughput 16S rRNA amplicon sequencing of the fecal samples was performed at the Life&Brain GmbH, Bonn, Germany. Sample transfer was done on dry ice. In brief, DNA was isolated from approximately 100 mg material of using ZR BashingBead lysis tubes (0.1 and 0.5 mm, Zymo Research, Freiburg, Germany) combined with the chemagic DNA Stool 200 Kit H96 (Perkin Elmer, Rodgau, Germany), according to the manufacturer’s instructions^131–133^. An additional mechanical lysis step was carried out by adding the lysis buffer and using the Precellys 24 Tissue Homogenizer (Bertin Instruments, Frankfurt am Main, Germany). After DNA extraction, the samples were stored at-20°C until further processing.

Details of library preparation and the 16S metagenomics analysis of the fecal microbiome have been published previously^134^. Briefly, the V3V4 region of the 16S rRNA gene was amplified using the primer combination Bakt_341F and Bakt_805R. Sequencing was performed on an Illumina MiSeq system using the MiSeq Reagent Kit v3 to generate 2 x 300 bp. Clustering was performed at 4 pM with a 15% spike-in of PhiX.

#### Genetic analysis

DNA was extracted from whole blood samples by a PerkinElmer chemagen System (Baesweiler, Germany) and magnetic bead-based extraction protocol and genotyped using llumina Inc. Global Screening Array (GSA) v3.0+MD (San Diego, CA, USA).

### Quantification and Statistical analysis

All statistical analyses were performed using IBM SPSS Statistics (v.29.0.1.1, IBM Corp., Chicago, IL, USA), R (versions 4.1.2 and 4.3.2, Boston, MA, USA) or GraphPad Prism for Mac (v.10.2.2, Boston, MA, USA). Univariate analyses (correlation and tests for differences as specified in the following chapters) were performed to investigate differences between RS and NRS in the top metabolic, genomic, lipidomic and microbial features as identified by sPLS-DA, DIABLO and XGBoost models in detail. All analyses were performed for the different diet groups individually, as well as in a combined approach, irrespective of the specific diet. Data are presented as mean ± standard error of the mean (SEM), unless stated otherwise. The significance level for all analyses was set at *p*<0.05. *p*-values are reported as *p*_unadj_ before correcting for multiple testing and as *p*_adj_ when adjusted. For the postprandial measurements (glucose, insulin, GLP-1, TAG and NEFA) the area under the curve (AUC) or incremental AUC (iAUC) were calculated using the integrated function in GraphPad Prism.

#### Pooling datasets

For the analyses, datasets from the first and second study phase were combined to increase the statistical power and to enable splitting up the data into training and test datasets for the prediction models. The time between participation in the two study phases was on average 19 months (9-30 months), ensuring a complete wash-out of intervention effects from the first study phase. A repetition of the screening visit with additional questions to assess if participants had changes in their habitual lifestyle or diet after participating in the first study phase was conducted, to ensure that the wash-out was completed, that participants returned to their initial lifestyle habits after participating in the first study and body weight stayed stable. To adjust for repeated participation and the two study phases, a sensitivity analysis was performed for the combined diet group, where only one observation (first study phase) was included for each participant to test the robustness of the results. Unless stated otherwise, results shown of the univariate statistical analyses of the all diets combined group have been confirmed by sensitivity analysis. In addition, linear-mixed model analyses were adjusted by adding a study variable as a covariate and using a common study ID.

### Metabolic dataset

Anthropometric measurements, resting energy expenditure, blood pressure and heart rate, the concentration of fecal butyric and acetic acid, the Shannon Diversity index, and a diet variable for analyses of the combined group, were integrated into the metabolic dataset. Variables that were represented by other variables, e.g., weight, height, waist and hip circumference by BMI, waist-height-ratio and waist-hip-ratio or fat free mass by fat mass, were removed from the dataset due to high correlation between them. Baseline characteristics between RS and NRS were compared using Student’s *t*-test for the diet groups and for all diets combined. For variables with a normal distribution, raw data was used. For other variables (insulinogenic and disposition index and zonulin), a Mann-Whitney-U-Test was used as a non-parametric alternative with log_10_(x+42) transformed data. Intervention effects were analyzed with linear mixed-models with responder status (RS or NRS), visit (baseline and endline) and responder status-visit interaction as fixed effects for each diet group (HD, ND, VD) and for all diets combined. The ID as identifier of the subjects was set as the random effect and a study variable classifying participation in the first or second study phase was set as covariate to control for possible effects of the two study phases. Hence, no additional sensitivity analysis was conducted for the results of the linear mixed-models. Prior to analysis, the data was log_10_(x+245) transformed and z-standardized if errors occurred. The residuals for all models were visually checked for notable deviations from normal distribution and homoscedasticity. All *p-*values were adjusted block-wise for multiple testing via Benjamini-Hochberg false discovery rate (FDR)^135^.

### Lipidomic data

Lipidomic data was log_10_(x+1) transformed before analysis. Differences between RS and NRS at baseline were assessed using linear regression models or zero-inflated gaussian models for each diet group (HD, ND, VD) and for all diets combined. The change in lipid species was examined using linear regression models where the change between endline and baseline was set as dependent variable, and baseline (and diet) as independent variables for all models. *P*-values were block-wise adjusted for the Benjamini-Hochberg FDR^135^. The change in lipid species is visualized using log 2-fold change (LFC) values.

### Microbiome data

Quality control of amplicon sequences, denoising, and joining were done in QIIME 2 version 2022.8^136^ using the DADA2 plugin^137^. The denoised sequences were classified using SILVA SSU 138 database to annotate amplicon sequencing variants (ASVs) for sequences with >99% sequence similarity. A rarefied table with a sampling depth of 69,770 reads was used to calculate alpha and beta diversity metrics.

Microbiome analyses were performed both at the Phylum and at the Genus taxonomic levels. For the analysis of differences between RS and NRS in diversity measures and taxonomy, we used filtered microbiome count data without CLR transformation. Relative abundance was computed for each sample-taxon pair by dividing a taxon’s count by the total sequencing depth of that sample. For each taxon and individual, the LFC in relative abundance between baseline and endline was calculated as the log₂ ratio of endline to baseline relative abundance, with a pseudocount of 10⁻□ added to avoid division by zero. To summarize group-level trends, relative abundance and LFC values were averaged across individuals within each diet and response group.

Differences in taxon-level relative abundance or in LFCs at baseline or endline were assessed between RS and NRS using the Wilcoxon rank-sum test. *P*-values were corrected for multiple comparisons using the Benjamini–Hochberg procedure to control the FDR^135^.

Microbial diversity was examined using alpha and beta diversity metrics. Alpha diversity was quantified using observed richness, Chao1 index, and Shannon index for all samples. Differences in alpha diversity metrics between RS and NRS groups were statistically evaluated within each dietary intervention and time point. First, the normality of the distribution of each alpha diversity metric across samples within each group was assessed using the Shapiro-Wilk normality test. When normality assumptions were met, a two-sample Student’s *t*-test was conducted; otherwise, a Wilcoxon rank-sum test was performed.

### Genetic data

In brief, polygenetic risk scores (PGS) were calculated using PRSice^138^ based on genome-wide association studies (GWAS) from the Global Lipids Genetics Consortium^139^ for TC, HDL-C and LDL-C and total TAG.

### Single-and multi-omics analyses

We obtained four omics-datasets from participants with two primary analytical objectives: predicting dietary response using baseline data and characterizing RS and NRS using baseline data and changes from baseline to endline. The input to the single-and multi-omics models that were calculated to address these questions consisted of four omics-datasets comprising different types of data as outlined above: metabolic data, PGS for hypercholesterinemia and TAG, 16S-derived metagenomics and lipidomic data. Each of the datasets was included as baseline or change data (from baseline to endline, except for PGS, as there is no change data).

#### Data preparation

Given the complexity and high dimensionality of multi-omics data, a detailed pre-processing approach was employed (**Figure S3 A-C**).

For predictive analyses, to avoid full rows of missing values within omics layers, we retained only participants with data available in all four omics blocks by identifying the set of common participants across datasets. The dataset was then split into training and test sets using a leave-one-out cross-validation (LOOCV) strategy. Accordingly, the following pre-processing steps were conducted separately for each train–test split. We applied an omics-based filtering approach, where lipidomics data retained lipid species with at least 5% of non-zero values in samples, while microbiome data included taxa detected in at least 1% of samples. LDL-C and TC were retained in the metabolic layer for the predictive models, as we wanted to capture the predictive power of these variables at baseline.

Metabolic and lipidomic datasets underwent log-transformation log_10_(x + c) to reduce skewness and stabilize variance, where c refers to the pseudocount used to avoid taking the logarithm of zero or negative values. For the metabolic dataset, the pseudocount was set slightly above the maximum negative value across all datasets (c = 42). For the lipidomics dataset, it was set to a positive value to avoid taking the logarithm of zero lipid concentrations (c = 1). Microbiome data was aggregated at the genus level and transformed using the Centered Log-Ratio (CLR)^140^. The CLR transformation was done using the *Microbiome* R Package (version 1.24.0), which applies a pseudocount to zero-count entries before taking the logarithm of those values^141^. For the PGS dataset, the PGS-TAG marker was previously transformed using logarithmic transformation. Density and QQ-plots indicated that the remaining PGS variables followed a normal distribution, thus they remained untransformed. Finally, features were harmonized across training and test sets by retaining only the common features present in both datasets after filtering (**Figure S3 B**).

For descriptive analyses, we used both baseline and change data. Different strategies for handling missing data were applied prior to pre-processing, depending on the specific model. For DIABLO, we included all available participants without restricting the blocks to retain common participants only, as the method demonstrated robustness to missing data. In contrast, sPLS-DA was more sensitive to missing values; thus, imputation was performed prior to model fitting. Imputation of missing values was performed on the metabolic, lipidomic, and PGS datasets after sub setting them by common identifiers, to avoid imputing rows with entirely missing omics data. Prior to imputation, only numerical variables were retained after excluding metadata, features with zero variance, and perfectly collinear variables. The imputation was carried out using the *mice* R package^142^ (version 3.17.0) with Predictive Mean Matching (PMM), and the predictor matrix was constructed using variables with a minimum absolute correlation of 0.2. A small ridge regularization term (ridge = 1e-2) was applied to enhance convergence in the presence of multicollinearity. Next, pre-processing filters and transformations were performed analogously to the prediction models, only this time using full datasets. Similar to the preprocessing for predictive models, the pseudocount for the metabolic dataset was set above the maximum negative value across both visits (c = 245), whereas for the lipidomics it was set to a positive value (c = 2). Baseline datasets were not pre-processed any further. Baseline and endline datasets were used to calculate changes from baseline to endline across all omics datasets. The change in metabolic markers was calculated from *log_10_*-transformed data as 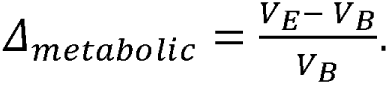 As the data was previously *log_10_*-transformed, a value of zero for *Δ*_metabolic_ reflects no change between baseline and endline, and *Δ*_metabolic_ can take negative values if the baseline value is greater than the endline value. On the other hand, changes in lipids and microbiome were calculated from log10-transformed and-CRL data, respectively. A change in these makers was calculated as 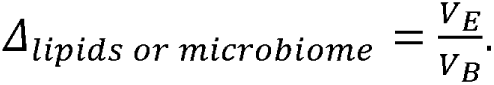 As the data was previously transformed, this quantity reflects a ratio of log values, where a value of 1 for *Δ*_lipids or microbiome_ reflects no change between baseline and endline values, and values below one represent a decrease from baseline to endline. Variables that did not vary between baseline and endline such as age, sex and all PGS markers were appended to the change datasets after the change computation (**Figure S3 C**).

#### Predictive models

EXtreme Gradient Boosting (XGBoost)^37^ (v. 1.7.10.1 R) was used to predict participants’ response to the different diets based on baseline multi-and single-omics data. Two primary approaches were used for fitting the models. On the one hand, we trained separate models for each dietary group to investigate response within a particular diet. On the other hand, all data was used to train models to predict response irrespective of the dietary intervention. In these models, we included diet-intervention-indicators to the metabolic data to account for the different interventions. Models were fit using a nested cross-validation approach. The outer loop used LOOCV, iteratively training on all but one participant, and validating the model on this held-out sample. Inside the inner loop, hyperparameter tunning was performed via 5-fold cross-validation (CV) using randomized search on 50 iterations. As shown in **Figure S3 B**, all four omics-datasets were taken into account to train the multi-omics models. Hyperparameters tuned included booster type (gradient boosting tree, DART^143^ or linear models), tree depth, learning rate, regularization parameters, subsampling rates, and other XGboost-specific parameters such as growth policy, sample type and feature normalization. Class imbalance was explicitly addressed by setting the ‘scale_pos_weight’ parameter to the RS and NRS frequencies observed in the train set. Performance was assessed using the performance metrics sensitivity, specificity, F1-score and AUC-ROC. Feature importance was systematically assessed and aggregated across LOOCV folds, separately for tree-based (gradient-boosting tree and DART) and linear models. Since tree-based models were by far the most frequently observed during training for the best performing models (**Table S4**), we only report tree-based aggregated performance metrics in the results section. Feature relevance was summarized using the gain metric and reported as mean gain, standard deviation, and frequency of appearance across folds.

To investigate the predictive power of each baseline omics dataset individually, we applied the same analytical framework described for the multi-omics analysis, restricting the input data to only one omics dataset at a time. Filtering and normalization procedures remained consistent with those previously described, but were adapted specifically for each omics type.

#### Descriptive models

For the identification of key discriminant features for RS from NRS, we trained ‘sparse Partial Least Squares – Discriminant Analysis (sPLS-DA)’ models from the mixOmics package^114^ (mixOmics v6.8.5 R) by projecting the data into a lower-dimensional space^144^. An initial model was evaluated via 5-fold CV to assess baseline performance. Hyperparameter tuning was done by repeated stratified 5-fold CV to select the optimal number of latent components and the number of variables retained per component which minimize the balanced error rate (BER). The maximum distance metric was applied to classify samples during CV by predicting the sample to the class with the highest predicted score value^144^. To get the optimal number of features retained per component, we kept 1 to 10 features and increased the subset of features in steps of 5 during hyperparameter tuning. For datasets with less than 10 features like PGS, all features were tested during CV. After this, a final model was re-fit using the optimal parameters and its performance was re-assessed via 5-fold CV using the BER and AUC metrics. The results are shown in sample plots, visualizing samples in a reduced dimensional space using the optimal number of components of the final model^114^. Sample plots are only reported for models with more than one optimal latent component, and include visualization of prediction areas for each class. For the sPLS-DA analyses using change data, LDL-C and TC (due to the strong correlation with LDL-C) were removed from the metabolic dataset to investigate the impact of other metabolic features characterizing RS. There were some special cases where the training strategy resulted in errors from the mixOmics package, especially the evaluation of the model’s performance using 5-fold CV. In these cases, the evaluation strategy was changed to either lower folds or LOOCV. Specifically, we used: 3-fold CV for the initial and last evaluation for the VD lipidomic baseline, VD metabolic change and VD lipidomic change analysis; 3-fold CV for the initial evaluation, inner tunning process and last evaluation for all diets combined PGS change analysis; LOOCV for the inner tunning process for ND lipidomic baseline analysis.

To investigate the relationship between metabolic, microbial, lipidomic, and PGS parameters and their potential in classifying RS and NRS, integrative multi-omics models were performed using “Data Integration Analysis for Biomarker discovery using Latent variable approaches for Omics studies (DIABLO)”^145^ (mixOmics v6.8.5 and v6.18.1 R). We first fitted an initial DIABLO model using no more than 4 components (as the PGS block has only four features) and stablishing the design matrix, which specifies the degree of the interaction between the different omics datasets. On a first iteration of the models, we tested different design settings (0, 0.25, 0.5, 0.75 and 1), where 0 reflects no correlation between blocks and 1 full correlation between blocks (**Table S7**). We determined the optimal number of components via repeated 5-fold-CV over 50 repeats using the maximum distance metric and evaluating the BER metric. Feature selection per component and omics layer was done in a second tuning step using repeated 5-fold-CV using maximum distance and the number of optimal components identified previously. A final model was then re-fitted using the design matrix, and the optimal number of latent components and features per block and per component. The prediction performance of the final model was assessed via LOOCV and the BER metric. We then took the design of the best performance model and fine-tuned the models again using two other distance metrics: centroids and mahalanobis distance (**Table S7**). While the maximum distance assigns a sample to the class with the highest predicted value, centroid-based distances consider the position of a sample relative to the class centroids in the latent component space. The mahalanobis distance additionally accounts for variance-covariance structure within each class. By assessing the model performance to other distances, we identified the most accurate classification for the DIABLO models (**Tables S7)**. For models with more than 1 optimal number of components, correlation coefficients were derived from pair-wise similarity matrixes from the mixOmics DIABLO models, which describe the correlation of the features with respect to the latent components, as well as correlation among features themselves^114,146^. We report block weights for each omics-layer and latent component, which reflect the correlation between each omics-layer and the outcome (RS, NRS) within each component^114^. The results of the DIABLO models are shown in circos and sample plots. In the circos plots, a cut-off value of 0.7 was used for visualizing correlations between features. Given the computational cost of performing 50 repetitions during hyperparameter tuning, we reduced the number of repetitions to 25 for the Mahalanobis distance in the all-diets combined change model and for all distance metrics in the all-diets combined baseline model.

#### Computational resources

HPC clusters were used for model fitting and evaluation of all DIABLO models and XGboost all diets combined multi-omics model. Specifically, the Marvin HPC cluster and IMBIE/IGSB HPC cluster from the University of Bonn were used. For DIABLO models, independent jobs were submitted for each distance metric and diet condition. Each job was allocated 20 CPU cores, 120 GB of memory, and a maximum running time of 7 days, and was executed in the intelsr long partition using R 4.3.2 (Marvin) and R 4.1.2 (IMBIE/IGSB). The average running time of each job was approximately 120 hours. All other models and analysis were executed locally on standard hardware.

## Additional Resources

The trial was prospectively registered at the German Clinical Trial Register (http://www.drks.de) under the identifier DRKS00015861.

## SUPPLEMENTAL INFORMATION

### Document Supplemental items

- Table S2
- Table S5
- Table S6
- Figures S1-S3

### Separate excel files

- **Table S1:** related to Fig. 2 – Fig. 6

⃘ Supplementary table S1-1: Performance characteristics of the best DIABLO models characterizing RS and NRS at baseline
⃘ Supplementary table S1-2: Performance characteristics of the best DIABLO models characterizing the change in multi-omics features of RS and NRS
⃘ Supplementary table S1-3: Performance characteristics and main results of the sPLS-DA models characterizing RS and NRS at baseline
⃘ Supplementary table S1-4: Performance characteristics and main results of the sPLS-DA models characterizing the change in features of RS and NRS
⃘ Supplementary table S1-5: Block weights showing the contribution of each single-omics dataset to the classification of RS and NRS in the DIABLO models
⃘ Supplementary table S1-6: Selected features and their weight loadings (importance) of the best DIABLO models characterizing RS and NRS at baseline
⃘ Supplementary table S1-7: Selected features and their weight loadings (importance) of the sPLS-DA models characterizing RS and NRS at baseline
⃘ Supplementary table S1-8: Selected features and their weight loadings (importance) of the best DIABLO models characterizing the change in multi-omics features of RS and NRS
⃘ Supplementary table S1-9: Selected features and their weight loadings (importance) of the sPLS-DA models characterizing the change in features of RS and NRS
- **Table S3:** related to Fig. 2 – Fig. 6

⃘ Supplementary table S3-1: Correlation matrix of the DIABLO model for all diets combined characterizing the change in multi-omics features of RS and NRS
⃘ Supplementary table S3-2: Correlation matrix of the DIABLO model for ND combined characterizing the change in multi-omics features of RS and NRS
⃘ Supplementary table S3-3: Correlation matrix of the DIABLO model for VD characterizing the change in multi-omics features of RS and NRS
⃘ Supplementary table S3-4: Correlation analysis (Pearson and Spearman’s rho) of the top features from the DIABLO (top 5 from each component) and XGBoost (top 10) models for all diets combined characterizing RS and NRS at baseline
⃘ Supplementary table S3-5: Correlation analysis (Pearson and Spearman’s rho) of the top features from the DIABLO (top 5 from each component) and XGBoost (top 10) models for ND characterizing RS and NRS at baseline
⃘ Supplementary table S3-6: Correlation analysis (Pearson and Spearman’s rho) of the top features from the DIABLO (top 5 from each component) and XGBoost (top 10) models for VD characterizing RS and NRS at baseline
⃘ Supplementary table S3-7: Correlation analysis (Pearson and Spearman’s rho) of the top features (top 5 from each component) from the best DIABLO model for all diets combined characterizing the change in multi-omics features of RS and NRS
⃘ Supplementary table S3-8: Correlation analysis (Pearson and Spearman’s rho) of the top features (top 5 from each component) from the best DIABLO model for ND characterizing characterizing the change in multi-omics features of RS and NRS
⃘ Supplementary table S3-9: Correlation analysis (Pearson and Spearman’s rho) of the top features (top 5 from each component) from the best DIABLO model for VD characterizing characterizing the change in multi-omics features of RS and NRS
- **Table S4:** related to Fig. 2 and Fig. 3 and to STAR methods

⃘ Supplementary table S4-1: Performance measures of the XGBoost single-and multi-omics prediction models
⃘ Supplementary table S4-2: Features and mean gain (importance) of the XGBoost single-and multi-omics prediction models
⃘ Supplementary table S4-3: Distribution of booster-types observed in XGBoost models
- **Table S7**, related to STAR methods

⃘ Supplementary table S7-1: Performance characteristics of DIABLO models with different design settings to characterize RS and NRS at baseline
⃘ Supplementary table S7-2: Performance characteristics of DIABLO models with different design settings to characterize the change in multi-omics features of RS and NRS
⃘ Supplementary table S7-3: Comparing the BER of different distance metrics (max, mahalanobis, centroids) for the best DIABLO models characterizing RS and NRS at baseline
⃘ Supplementary table S7-4: Comparing the BER of different distance metrics (max, mahalanobis, centroids) for the best DIABLO models characterizing the change in multi-omics features of RS and NRS

## References

1. Kim, J. Y. Optimal Diet Strategies for Weight Loss and Weight Loss Maintenance. J. Obes. Metab. Syndr. 30, 20–31 (2021).

2. Aaseth, J., Ellefsen, S., Alehagen, U., Sundfør, T. M. & Alexander, J. Diets and drugs for weight loss and health in obesity – An update. Biomed. Pharmacother. 140, 111789 (2021).

3. Muscogiuri, G. et al. European Guidelines for Obesity Management in Adults with a Very Low-Calorie Ketogenic Diet: A Systematic Review and Meta-Analysis. Obes. Facts 14, 222–245 (2021).

4. Asnicar, F. et al. Microbiome connections with host metabolism and habitual diet from 1,098 deeply phenotyped individuals. Nat. Med. 27, 321–332 (2021).

5. Zeevi, D. et al. Personalized Nutrition by Prediction of Glycemic Responses. Cell 163, 1079–1094 (2015).

6. Berry, S. E. et al. Human postprandial responses to food and potential for precision nutrition. Nat. Med. 26, 964–973 (2020).

7. Palmnäs, M. et al. Perspective: Metabotyping—A Potential Personalized Nutrition Strategy for Precision Prevention of Cardiometabolic Disease. Adv. Nutr. 11, 524– 532 (2020).

8. Simon, M., Sina, C., Ferrario, P. G., Daniel, H., & Working Group “Personalized Nutrition” of the German Nutrition Society. Gut Microbiome Analysis for Personalized Nutrition: The State of Science. Mol. Nutr. Food Res. 67, 2200476 (2023).

9. Holzapfel, C., Waldenberger, M., Lorkowski, S., Daniel, H., & the Working Group “Personalized Nutrition” of the German Nutrition Society. Genetics and Epigenetics in Personalized Nutrition: Evidence, Expectations, and Experiences. Mol. Nutr. Food Res. 66, 2200077 (2022).

10. Kirwan, L. et al. Phenotypic factors influencing the variation in response of circulating cholesterol level to personalised dietary advice in the Food4Me study. Br. J. Nutr. 116, 2011–2019 (2016).

11. Korpela, K. et al. Gut Microbiota Signatures Predict Host and Microbiota Responses to Dietary Interventions in Obese Individuals. PLoS ONE 9, e90702 (2014).

12. Lauber, C. et al. Lipidomic risk scores are independent of polygenic risk scores and can predict incidence of diabetes and cardiovascular disease in a large population cohort. PLOS Biol. 20, e3001561 (2022).

13. Bosak, K., Sauer, A. & Meeusen, J. Clinical Update: Ceramides As Novel Biomarkers of Cardiovascular Disease Risk. J. Nurse Pract. 20, 104838 (2024).

14. Li, Y. et al. Lipidomics identified novel cholesterol-independent predictors for risk of incident coronary heart disease: Mediation of risk from diabetes and aggravation of risk by ambient air pollution. J. Adv. Res. S209012322300396X (2023) doi:10.1016/j.jare.2023.12.009.

15. Bellot, P. E. N. R. et al. Plasma lipid metabolites as potential biomarkers for identifying individuals at risk of obesity-induced metabolic complications. Sci. Rep. 13, 11729 (2023).

16. Eichelmann, F. et al. Lipidome changes due to improved dietary fat quality inform cardiometabolic risk reduction and precision nutrition. Nat. Med. 1–11 (2024) doi:10.1038/s41591-024-03124-1.

17. Ambegaonkar, B. M. et al. Attainment of normal lipid levels among high cardiovascular risk patients: Pooled analysis of observational studies from the United Kingdom, Sweden, Spain and Canada. Eur. J. Intern. Med. 24, 656–663 (2013).

18. Ajala, O., English, P. & Pinkney, J. Systematic review and meta-analysis of different dietary approaches to the management of type 2 diabetes. Am. J. Clin. Nutr. 97, 505–516 (2013).

19. Bahls, M. et al. Progression of conventional cardiovascular risk factors and vascular disease risk in individuals: insights from the PROG-IMT consortium. Eur. J. Prev. Cardiol. 27, 234–243 (2020).

20. Goldberg, R. B. et al. Effect of Progression From Impaired Glucose Tolerance to Diabetes on Cardiovascular Risk Factors and Its Amelioration by Lifestyle and Metformin Intervention. Diabetes Care 32, 726–732 (2009).

21. Orozco-Beltran, D. et al. Lipid profile, cardiovascular disease and mortality in a Mediterranean high-risk population: The ESCARVAL-RISK study. PLOS ONE 12, e0186196 (2017).

22. Ference, B. A. et al. Low-density lipoproteins cause atherosclerotic cardiovascular disease. 1. Evidence from genetic, epidemiologic, and clinical studies. A consensus statement from the European Atherosclerosis Society Consensus Panel. Eur. Heart J. 38, 2459–2472 (2017).

23. Silverman, M. G. et al. Association Between Lowering LDL-C and Cardiovascular Risk Reduction Among Different Therapeutic Interventions: A Systematic Review and Meta-analysis. JAMA 316, 1289 (2016).

24. Wang, N. et al. Intensive LDL cholesterol-lowering treatment beyond current recommendations for the prevention of major vascular events: a systematic review and meta-analysis of randomised trials including 327 037 participants. Lancet Diabetes Endocrinol. 8, 36–49 (2020).

25. Cholesterol Treatment Trialists’ (Ctt) Collaborators. The effects of lowering LDL cholesterol with statin therapy in people at low risk of vascular disease: meta-analysis of individual data from 27 randomised trials. The Lancet 380, 581–590 (2012).

26. Wong, H. J. et al. Efficacy of GLP-1 Receptor Agonists on Weight Loss, BMI, and Waist Circumference for Patients With Obesity or Overweight: A Systematic Review, Meta-analysis, and Meta-regression of 47 Randomized Controlled Trials. Diabetes Care 48, 292–300 (2025).

27. Badve, S. V. et al. Effects of GLP-1 receptor agonists on kidney and cardiovascular disease outcomes: a meta-analysis of randomised controlled trials. Lancet Diabetes Endocrinol. 13, 15–28 (2025).

28. Burger, P. M. et al. Course of the effects of LDL-cholesterol reduction on cardiovascular risk over time: A meta-analysis of 60 randomized controlled trials. Atherosclerosis 396, (2024).

29. Capodici, A. et al. Cardiovascular health and cancer risk associated with plant based diets: An umbrella review. PLOS ONE 19, e0300711 (2024).

30. Aziz, T., Hussain, N., Hameed, Z. & Lin, L. Elucidating the role of diet in maintaining gut health to reduce the risk of obesity, cardiovascular and other age-related inflammatory diseases: recent challenges and future recommendations. Gut Microbes 16, 2297864 (2024).

31. Ramezani-Jolfaie, N., Mohammadi, M. & Salehi-Abargouei, A. The effect of healthy Nordic diet on cardio-metabolic markers: a systematic review and meta-analysis of randomized controlled clinical trials. Eur. J. Nutr. 58, 2159–2174 (2019).

32. Pigsborg, K. et al. Effects of changing from a diet with saturated fat to a diet with n-6 polyunsaturated fat on the serum metabolome in relation to cardiovascular disease risk factors. Eur. J. Nutr. 61, 2079–2089 (2022).

33. Melgar, B. et al. Vegetarian diets on anthropometric, metabolic and blood pressure outcomes in people with overweight and obesity: a systematic review and meta-analysis of randomized controlled trials. Int. J. Obes. 47, 903–910 (2023).

34. Oussalah, A., Levy, J., Berthezène, C., Alpers, D. H. & Guéant, J.-L. Health outcomes associated with vegetarian diets: An umbrella review of systematic reviews and meta-analyses. Clin. Nutr. 39, 3283–3307 (2020).

35. Massara, P. et al. Nordic dietary patterns and cardiometabolic outcomes: a systematic review and meta-analysis of prospective cohort studies and randomised controlled trials. Diabetologia 65, 2011–2031 (2022).

36. Clemente-Suárez, V. J., Beltrán-Velasco, A. I., Redondo-Flórez, L., Martín-Rodríguez, A. & Tornero-Aguilera, J. F. Global Impacts of Western Diet and Its Effects on Metabolism and Health: A Narrative Review. Nutrients 15, 2749 (2023).

37. Chen, T. & Guestrin, C. XGBoost: A Scalable Tree Boosting System. in Proceedings of the 22nd ACM SIGKDD International Conference on Knowledge Discovery and Data Mining 785–794 (Association for Computing Machinery, New York, NY, USA, 2016). doi:10.1145/2939672.2939785.

38. Bu, T., Sun, Z., Pan, Y., Deng, X. & Yuan, G. Glucagon-Like Peptide-1: New Regulator in Lipid Metabolism. Diabetes Metab. J. 48, 354–372 (2024).

39. Mehmood, A. et al. Evaluation of Lipid Profile Alterations for Early Diagnosis and Therapeutic Management of Cardiovascular and Metabolic Disorders. Pak. J. Med. Health Sci. 17, 53–53 (2023).

40. Jung, E., Kong, S. Y., Ro, Y. S., Ryu, H. H. & Shin, S. D. Serum Cholesterol Levels and Risk of Cardiovascular Death: A Systematic Review and a Dose-Response Meta-Analysis of Prospective Cohort Studies. Int. J. Environ. Res. Public. Health 19, 8272 (2022).

41. Rämö, J. T. et al. Coronary artery disease risk and lipidomic profiles are similar in familial and population-ascertained hyperlipidemias. 321752 Preprint at 10.1101/321752 (2018).

42. Navarese, E. P. et al. Association Between Baseline LDL-C Level and Total and Cardiovascular Mortality After LDL-C Lowering: A Systematic Review and Meta-analysis. JAMA 319, 1566 (2018).

43. Marston, N. A. et al. Association Between Triglyceride Lowering and Reduction of Cardiovascular Risk Across Multiple Lipid-Lowering Therapeutic Classes. Circulation 140, 1308–1317 (2019).

44. Cholesterol Treatment Trialists’ (Ctt) Collaboration. Efficacy and safety of more intensive lowering of LDL cholesterol: a meta-analysis of data from 170 000 participants in 26 randomised trials. The Lancet 376, 1670–1681 (2010).

45. Massini, G. et al. Mediterranean Dietary Treatment in Hyperlipidemic Children: Should It Be an Option? Nutrients 14, 1344 (2022).

46. Polygenic Risk Score: Clinically Useful Tool for Prediction of Cardiovascular Disease and Benefit from Lipid-Lowering Therapy? | Cardiovascular Drugs and Therapy. https://link.springer.com/article/10.1007/s10557-020-07105-7#Sec3.

47. Khera, A. V. et al. Genetic Risk, Adherence to a Healthy Lifestyle, and Coronary Disease. N. Engl. J. Med. 375, 2349–2358 (2016).

48. Dron, J. S. The clinical utility of polygenic risk scores for combined hyperlipidemia. Curr. Opin. Lipidol. 34, 44 (2023).

49. Huber, H. et al. Differential effects of Nordic and Vegetarian diets on lipid metabolism, gut microbiome and cardiometabolic risk factors: A multi-omic perspective from a randomized clinical intervention trial. 2025.11.03.25337885 Preprint at 10.1101/2025.11.03.25337885 (2025).

50. Sánchez-Archidona, A. R. et al. Plasma triacylglycerols are biomarkers of β-cell function in mice and humans. Mol. Metab. 54, 101355 (2021).

51. Hornburg, D. et al. Dynamic lipidome alterations associated with human health, disease and ageing. Nat. Metab. 5, 1578–1594 (2023).

52. Toledo, E. et al. Plasma lipidomic profiles and cardiovascular events in a randomized intervention trial with the Mediterranean diet. Am. J. Clin. Nutr. 106, 973–983 (2017).

53. Vanni, S., Riccardi, L., Palermo, G. & De Vivo, M. Structure and Dynamics of the Acyl Chains in the Membrane Trafficking and Enzymatic Processing of Lipids. Acc. Chem. Res. 52, 3087–3096 (2019).

54. Shi, H. et al. Effects of Pomegranate Peel Polyphenols Combined with Inulin on Gut Microbiota and Serum Metabolites of High-Fat-Induced Obesity Rats. J. Agric. Food Chem. 71, 5733–5744 (2023).

55. Acciarino, A. et al. The role of the gastrointestinal barrier in obesity-associated systemic inflammation. Obes. Rev. 25, e13673 (2024).

56. Vekic, J., Stefanovic, A. & Zeljkovic, A. Obesity and Dyslipidemia: A Review of Current Evidence. Curr. Obes. Rep. 12, 207–222 (2023).

57. Nussbaumerova, B. & Rosolova, H. Obesity and Dyslipidemia. Curr. Atheroscler. Rep. 25, 947–955 (2023).

58. Li, F. et al. Serum Uric Acid Levels and Metabolic Indices in an Obese Population: A Cross-Sectional Study. Diabetes Metab. Syndr. Obes. Targets Ther. 14, 627–635 (2021).

59. Zhang, S. Y. et al. Overweight, resting heart rate and prediabetes/diabetes: A population-based prospective cohort study among Inner Mongolians in China. Sci. Rep. 6, 23939 (2016).

60. Ahmed, B., Sultana, R. & Greene, M. W. Adipose tissue and insulin resistance in obese. Biomed. Pharmacother. 137, 111315 (2021).

61. Barber, T. M., Kyrou, I., Randeva, H. S. & Weickert, M. O. Mechanisms of Insulin Resistance at the Crossroad of Obesity with Associated Metabolic Abnormalities and Cognitive Dysfunction. Int. J. Mol. Sci. 22, 546 (2021).

62. Meldrum, D. R., Morris, M. A. & Gambone, J. C. Obesity pandemic: causes, consequences, and solutions—but do we have the will? Fertil. Steril. 107, 833–839 (2017).

63. Howard, B. V. Insulin resistance and lipid metabolism. Am. J. Cardiol. 84, 28–32 (1999).

64. Hoenig, M. R. & Sellke, F. W. Insulin resistance is associated with increased cholesterol synthesis, decreased cholesterol absorption and enhanced lipid response to statin therapy. Atherosclerosis 211, 260–265 (2010).

65. Oduncu, E. A., Tural, E. & Dayan, A. Evaluation of the relationship between oral glucose tolerance test, blood sugar levels and cholesterol levels in prediabetic individuals. Pak. J. Med. Sci. 40, (2024).

66. Bianchi, C. et al. Elevated 1-Hour Postload Plasma Glucose Levels Identify Subjects With Normal Glucose Tolerance but Impaired β-Cell Function, Insulin Resistance, and Worse Cardiovascular Risk Profile: The GENFIEV Study. J. Clin. Endocrinol. Metab. 98, 2100–2105 (2013).

67. Andreozzi, F., Mannino, G. C., Perticone, M., Perticone, F. & Sesti, G. Elevated 1-h post-load plasma glucose levels in subjects with normal glucose tolerance are associated with a pro-atherogenic lipid profile. Atherosclerosis 256, 15–20 (2017).

68. Ogita, K. et al. Serum concentration of small dense low-density lipoprotein-cholesterol during oral glucose tolerance test and oral fat tolerance test. Clin. Chim. Acta 387, 36–41 (2008).

69. Roden, M. et al. Mechanism of free fatty acid-induced insulin resistance in humans. J. Clin. Invest. 97, 2859–2865 (1996).

70. Roden, M. et al. Rapid impairment of skeletal muscle glucose transport/phosphorylation by free fatty acids in humans. Diabetes 48, 358–364 (1999).

71. Elkanawati, R. Y., Sumiwi, S. A. & Levita, J. Impact of Lipids on Insulin Resistance: Insights from Human and Animal Studies. Drug Des. Devel. Ther. 18, 3337–3360 (2024).

72. Trouwborst, I. et al. Cardiometabolic health improvements upon dietary intervention are driven by tissue-specific insulin resistance phenotype: A precision nutrition trial. Cell Metab. 35, 71–83.e5 (2023).

73. Blanco-Rojo, R. et al. The insulin resistance phenotype (muscle or liver) interacts with the type of diet to determine changes in disposition index after 2 years of intervention: the CORDIOPREV-DIAB randomised clinical trial. Diabetologia 59, 67– 76 (2016).

74. Nowotny, B. et al. Mechanisms Underlying the Onset of Oral Lipid–Induced Skeletal Muscle Insulin Resistance in Humans. Diabetes 62, 2240–2248 (2013).

75. Joshi, A., Rienks, M., Theofilatos, K. & Mayr, M. Systems biology in cardiovascular disease: a multiomics approach. Nat. Rev. Cardiol. 18, 313–330 (2021).

76. Knoll, R. et al. The life-saving benefit of dexamethasone in severe COVID-19 is linked to a reversal of monocyte dysregulation. Cell 187, 4318–4335.e20 (2024).

77. Wang, R., Li, B., Lam, S. M. & Shui, G. Integration of lipidomics and metabolomics for in-depth understanding of cellular mechanism and disease progression. J. Genet. Genomics 47, 69–83 (2020).

78. Livingstone, K. M. et al. Precision nutrition: A review of current approaches and future endeavors. Trends Food Sci. Technol. 128, 253–264 (2022).

79. Zhang, S., Niu, H. & Zhu, J. Personalized nutrition studies of human gut microbiome-polyphenol interactions utilizing continuous multistaged *in vitro* fermentation models– a narrative review. Nutr. Res. 135, 101–127 (2025).

80. Abuqwider, J. N., Mauriello, G. & Altamimi, M. Akkermansia muciniphila, a New Generation of Beneficial Microbiota in Modulating Obesity: A Systematic Review. Microorganisms 9, 1098 (2021).

81. Ashrafian, F. et al. Comparative effects of alive and pasteurized Akkermansia muciniphila on normal diet-fed mice. Sci. Rep. 11, 17898 (2021).

82. Depommier, C. et al. Supplementation with Akkermansia muciniphila in overweight and obese human volunteers: a proof-of-concept exploratory study. Nat. Med. 25, 1096–1103 (2019).

83. Salomaa, V. et al. Fatty acid composition of serum cholesterol esters in different degrees of glucose intolerance: A population-based study. Metabolism 39, 1285– 1291 (1990).

84. Eisinger, K. et al. Lipidomic Analysis of Serum from High Fat Diet Induced Obese Mice. Int. J. Mol. Sci. 15, 2991–3002 (2014).

85. Salamone, D. et al. Fatty acid composition of cholesterol esters reflects dietary fat intake after dietary interventions in a multinational population. J. Clin. Lipidol. 17, 466–474 (2023).

86. Huang, Y. et al. Lipid profiling identifies modifiable signatures of cardiometabolic risk in children and adolescents with obesity. Nat. Med. 31, 294–305 (2025).

87. Structure and Dynamics of the Acyl Chains in the Membrane Trafficking and Enzymatic Processing of Lipids | Accounts of Chemical Research. https://pubs.acs.org/doi/10.1021/acs.accounts.9b00134.

88. Li, C. et al. Gut microbiome and metabolome profiling in Framingham heart study reveals cholesterol-metabolizing bacteria. Cell 187, 1834–1852.e19 (2024).

89. Yan, S., et al. Oryzanol alleviates high fat and cholesterol diet-induced hypercholesterolemia associated with the modulation of the gut microbiota in hamsters. (2022).

90. Simon, M.-C., et al. Cholesterol-lowering effects of oats induced by microbially produced phenolic metabolites. Preprint at 10.21203/rs.3.rs-4188074/v2 (2024).

91. Määttä-Riihinen, K. R., Kamal-Eldin, A., Mattila, P. H., González-Paramás, A. M. & Törrönen, A. R. Distribution and Contents of Phenolic Compounds in Eighteen Scandinavian Berry Species. J. Agric. Food Chem. 52, 4477–4486 (2004).

92. Chen, C. et al. Phenolic contents, cellular antioxidant activity and antiproliferative capacity of different varieties of oats. Food Chem. 239, 260–267 (2018).

93. Yang, F. & Chen, G. The nutritional functions of dietary sphingomyelin and its applications in food. Front. Nutr. 9, (2022).

94. Mokkala, K., Houttu, N., Cansev, T. & Laitinen, K. Interactions of dietary fat with the gut microbiota: Evaluation of mechanisms and metabolic consequences. Clin. Nutr. 39, 994–1018 (2020).

95. Chadaideh, K. S. & Carmody, R. N. Host-microbial interactions in the metabolism of different dietary fats. Cell Metab. 33, 857–872 (2021).

96. Craig, W. J. Nutrition Concerns and Health Effects of Vegetarian Diets. Nutr. Clin. Pract. 25, 613–620 (2010).

97. Mithril, C. et al. Dietary composition and nutrient content of the New Nordic Diet. Public Health Nutr. 16, 777–785 (2013).

98. Sellem, L. et al. Replacement of dietary saturated with unsaturated fatty acids is associated with beneficial effects on lipidome metabolites: a secondary analysis of a randomized trial. Am. J. Clin. Nutr. 117, 1248–1261 (2023).

99. Schwingshackl, L. et al. Total Dietary Fat Intake, Fat Quality, and Health Outcomes: A Scoping Review of Systematic Reviews of Prospective Studies. Ann. Nutr. Metab. 77, 4–15 (2021).

100. Gao, Y., Xun, R., Xia, J., Xia, H. & Sun, G. Effects of phytosterol supplementation on lipid profiles in patients with hypercholesterolemia: a systematic review and meta-analysis of randomized controlled trials. Food Funct. 14, 2969–2997 (2023).

101. Ghavami, A. et al. Soluble Fiber Supplementation and Serum Lipid Profile: A Systematic Review and Dose-Response Meta-Analysis of Randomized Controlled Trials. Adv. Nutr. 14, 465–474 (2023).

102. Kalupahana, N. S., Claycombe, K. J. & Moustaid-Moussa, N. (n-3) Fatty acids alleviate adipose tissue inflammation and insulin resistance: mechanistic insights. Adv. Nutr. Bethesda Md 2, 304–316 (2011).

103. Chacińska, M. et al. The Impact of OMEGA-3 Fatty Acids Supplementation on Insulin Resistance and Content of Adipocytokines and Biologically Active Lipids in Adipose Tissue of High-Fat Diet Fed Rats. Nutrients 11, 835 (2019).

104. Bunge, A. C., Mazac, R., Clark, M., Wood, A. & Gordon, L. Sustainability benefits of transitioning from current diets to plant-based alternatives or whole-food diets in Sweden. Nat. Commun. 15, 951 (2024).

105. Carey, C. N. et al. The Environmental Sustainability of Plant-Based Dietary Patterns: A Scoping Review. J. Nutr. 153, 857–869 (2023).

106. Trolle, E. et al. Integrating environmental sustainability into food-based dietary guidelines in the Nordic countries. Food Nutr. Res. 10.29219/fnr.v68.10792 (2024) doi:10.29219/fnr.v68.10792.

107. Schulz, C.-A., Oluwagbemigun, K. & Nöthlings, U. Advances in dietary pattern analysis in nutritional epidemiology. Eur. J. Nutr. 60, 4115–4130 (2021).

108. Hjorth, T. et al. Effectiveness of regular oat β-glucan–enriched bread compared with whole-grain wheat bread on long-term glycemic control in adults at risk of type 2 diabetes: a randomized controlled trial. Am. J. Clin. Nutr. 10.1016/j.ajcnut.2025.06.018(2025) doi:10.1016/j.ajcnut.2025.06.018.

109. Viroli, G., Kalmpourtzidou, A. & Cena, H. Exploring Benefits and Barriers of Plant-Based Diets: Health, Environmental Impact, Food Accessibility and Acceptability. Nutrients 15, 4723 (2023).

110. Mithril, C. et al. Guidelines for the New Nordic Diet. Public Health Nutr. 15, 1941– 1947 (2012).

111. Nutrition research must go local. Nat. Med. 31, 1371–1371 (2025).

112. Stratakis, N. et al. Multi-omics architecture of childhood obesity and metabolic dysfunction uncovers biological pathways and prenatal determinants. Nat. Commun. 16, 654 (2025).

113. Performance assessment and parameter tuning – mixOmics. https://mixomics.org/performance-assessment-and-parameter-tuning/.

114. Rohart, F., Gautier, B., Singh, A. & Cao, K.-A. L. mixOmics: An R package for ‘omics feature selection and multiple data integration. PLOS Comput. Biol. 13, e1005752 (2017).

115. World Health Organization. Definition, Diagnosis and Classification of Diabetes Mellitus and Its Complications: Report of a WHO Consultation. Part 1: Diagnosis and Classification of Diabetes Mellitus. (1999).

116. Hanna Huber. Dissertation: Impact of Dietary Interventions on Metabolism and the Gut-Brain Axis in Adults with Metabolic Syndrome Traits. (2023).

117. Peng, K., Li, X., Wang, Z., Li, M. & Yang, Y. Association of low-density lipoprotein cholesterol levels with the risk of mortality and cardiovascular events: A meta-analysis of cohort studies with 1,232,694 participants. Medicine (Baltimore) 101, e32003 (2022).

118. Poulsen, S. K. et al. Health effect of the New Nordic Diet in adults with increased waist circumference: a 6-mo randomized controlled trial. Am. J. Clin. Nutr. 99, 35–45 (2014).

119. Adamsson, V. et al. Effects of a healthy Nordic diet on cardiovascular risk factors in hypercholesterolaemic subjects: a randomized controlled trial (NORDIET): Diet and cardiovascular risk factors. J. Intern. Med. 269, 150–159 (2011).

120. Ohlsson, B. An OkinawanlZbased Nordic diet improves glucose and lipid metabolism in health and type 2 diabetes, in alignment with changes in the endocrine profile, whereas zonulin levels are elevated (Review). Exp. Ther. Med. 10.3892/etm.2019.7303(2019) doi:10.3892/etm.2019.7303.

121. Uusitupa, M. et al. Effects of an isocaloric healthy N ordic diet on insulin sensitivity, lipid profile and inflammation markers in metabolic syndrome – a randomized study (SYSDIET). J. Intern. Med. 274, 52–66 (2013).

122. Wang, F. et al. Effects of Vegetarian Diets on Blood Lipids: A Systematic Review and Meta-Analysis of Randomized Controlled Trials. J. Am. Heart Assoc. 4, e002408 (2015).

123. Yokoyama, Y., Levin, S. M. & Barnard, N. D. Association between plant-based diets and plasma lipids: a systematic review and meta-analysis. Nutr. Rev. 75, 683–698 (2017).

124. Feingold, K. R. Cholesterol Lowering Drugs. Endotext (2000).

125. Penson, P. E., Pirro, M. & Banach, M. LDL-C: lower is better for longer—even at low risk. BMC Med. 18, 320 (2020).

126. Bak, M. J. et al. Specificity and sensitivity of commercially available assays for glucagon-like peptide-1 (GLP-1): implications for GLP-1 measurements in clinical studies. Diabetes Obes. Metab. 16, 1155–1164 (2014).

127. Matthews, D. R. et al. Homeostasis model assessment: insulin resistance and beta-cell function from fasting plasma glucose and insulin concentrations in man. Diabetologia 28, 412–419 (1985).

128. Gutch, M., Kumar, S., Razi, S. M., Gupta, K. K. & Gupta, A. Assessment of insulin sensitivity/resistance. Indian J. Endocrinol. Metab. 19, 160 (2015).

129. Patarrão, R. S., Wayne Lautt, W. & Paula Macedo, M. Assessment of methods and indexes of insulin sensitivity. Rev. Port. Endocrinol. Diabetes E Metab. 9, 65–73 (2014).

130. Mari, A., Pacini, G., Murphy, E., Ludvik, B. & Nolan, J. J. A model-based method for assessing insulin sensitivity from the oral glucose tolerance test. Diabetes Care 24, 539–548 (2001).

131. Junkins, E. N., McWhirter, J. B., McCall, L.-I. & Stevenson, B. S. Environmental structure impacts microbial composition and secondary metabolism. ISME Commun. 2, 1–10 (2022).

132. Kable, M. E., Chin, E. L., Storms, D., Lemay, D. G. & Stephensen, C. B. Tree-Based Analysis of Dietary Diversity Captures Associations Between Fiber Intake and Gut Microbiota Composition in a Healthy US Adult Cohort. J. Nutr. 152, 779–788 (2022).

133. Neuberger-Castillo, L. et al. Method Validation for Extraction of DNA from Human Stool Samples for Downstream Microbiome Analysis. Biopreservation Biobanking 18, 102–116 (2020).

134. Seel, W., Reiners, S., Kipp, K., Simon, M.-C. & Dawczynski, C. Role of Dietary Fiber and Energy Intake on Gut Microbiome in Vegans, Vegetarians, and Flexitarians in Comparison to Omnivores—Insights from the Nutritional Evaluation (NuEva) Study. Nutrients 15, 1914 (2023).

135. Benjamini, Y. & Hochberg, Y. Controlling the False Discovery Rate: A Practical and Powerful Approach to Multiple Testing. J. R. Stat. Soc. Ser. B Methodol. 57, 289–300 (1995).

136. Bolyen, E. et al. Reproducible, interactive, scalable and extensible microbiome data science using QIIME 2. Nat. Biotechnol. 37, 852–857 (2019).

137. Callahan, B. J. et al. DADA2: High-resolution sample inference from Illumina amplicon data. Nat. Methods 13, 581–583 (2016).

138. Euesden, J., Lewis, C. M. & O’Reilly, P. F. PRSice: Polygenic Risk Score software. Bioinformatics 31, 1466–1468 (2015).

139. Teslovich, T. M. et al. Biological, clinical and population relevance of 95 loci for blood lipids. Nature 466, 707–713 (2010).

140. Gloor, G. B., Macklaim, J. M., Pawlowsky-Glahn, V. & Egozcue, J. J. Microbiome Datasets Are Compositional: And This Is Not Optional. Front. Microbiol. 8, 2224 (2017).

141. Introduction to the microbiome R package. https://microbiome.github.io/tutorials/.

142. Buuren, S. van & Groothuis-Oudshoorn, K. mice: Multivariate Imputation by Chained Equations in R. J. Stat. Softw. 45, 1–67 (2011).

143. Vinayak, R. K. & Gilad-Bachrach, R. DART: Dropouts meet Multiple Additive Regression Trees. in Proceedings of the Eighteenth International Conference on Artificial Intelligence and Statistics 489–497 (PMLR, 2015).

144. Lê Cao, K.-A., Boitard, S. & Besse, P. Sparse PLS discriminant analysis: biologically relevant feature selection and graphical displays for multiclass problems. BMC Bioinformatics 12, 253 (2011).

145. Singh, A. et al. DIABLO: an integrative approach for identifying key molecular drivers from multi-omics assays. Bioinforma. Oxf. Engl. 35, 3055–3062 (2019).

146. González, I., Cao, K.-A. L., Davis, M. J. & Déjean, S. Visualising associations between paired ‘omics’ data sets. BioData Min. 5, 19 (2012).

