## Supplemental Items for "Identifying Multi-omics Signatures that characterize Responders to Plant-based Dietary Interventions"

**Tables:**

- **Table S1:** provided in separate excel file, related to Fig. 2 – Fig. 6
  - Supplementary table S1-1: Performance characteristics of the best DIABLO models characterizing RS and NRS at baseline
  - Supplementary table S1-2: Performance characteristics of the best DIABLO models characterizing the change in multi-omics features of RS and NRS
  - Supplementary table S1-3: Performance characteristics and main results of the sPLS-DA models characterizing RS and NRS at baseline
  - Supplementary table S1-4: Performance characteristics and main results of the sPLS-DA models characterizing the change in features of RS and NRS
  - Supplementary table S1-5: Block weights showing the contribution of each single-omics dataset to the classification of RS and NRS in the DIABLO models
  - Supplementary table S1-6: Selected features and their weight loadings (importance) of the best DIABLO models characterizing RS and NRS at baseline
  - Supplementary table S1-7: Selected features and their weight loadings (importance) of the sPLS-DA models characterizing RS and NRS at baseline
  - Supplementary table S1-8: Selected features and their weight loadings (importance) of the best DIABLO models characterizing the change in multi-omics features of RS and NRS
  - Supplementary table S1-9: Selected features and their weight loadings (importance) of the sPLS-DA models characterizing the change in features of RS and NRS
- **Table S2:** Additional baseline characteristics of RS, NRS and all participants in the different diet groups, related to Fig. 2 and Fig. 3
- **Table S3:** provided in separate excel file, related to Fig. 2 – Fig. 6
  - Supplementary table S3-1: Correlation matrix of the DIABLO model for all diets combined characterizing the change in multi-omics features of RS and NRS
  - Supplementary table S3-2: Correlation matrix of the DIABLO model for ND combined characterizing the change in multi-omics features of RS and NRS
  - Supplementary table S3-3: Correlation matrix of the DIABLO model for VD characterizing the change in multi-omics features of RS and NRS
  - Supplementary table S3-4: Correlation analysis (Pearson and Spearman's rho) of the top features from the DIABLO (top 5 from each component) and XGBoost (top 10) models for all diets combined characterizing RS and NRS at baseline
  - Supplementary table S3-5: Correlation analysis (Pearson and Spearman's rho) of the top features from the DIABLO (top 5 from each component) and XGBoost (top 10) models for ND characterizing RS and NRS at baseline
  - Supplementary table S3-6: Correlation analysis (Pearson and Spearman's rho) of the top features from the DIABLO (top 5 from each component) and XGBoost (top 10) models for VD characterizing RS and NRS at baseline
  - Supplementary table S3-7: Correlation analysis (Pearson and Spearman's rho) of the top features (top 5 from each component) from the best DIABLO model for all diets combined characterizing the change in multi-omics features of RS and NRS
  - Supplementary table S3-8: Correlation analysis (Pearson and Spearman's rho) of the top features (top 5 from each component) from the best DIABLO model for ND characterizing characterizing the change in multi-omics features of RS and NRS
  - Supplementary table S3-9: Correlation analysis (Pearson and Spearman's rho) of the top features (top 5 from each component) from the best DIABLO model for VD characterizing characterizing the change in multi-omics features of RS and NRS
- **Table S4:** provided in separate excel file, related to Fig. 2 and Fig. 3 and to STAR methods
  - Supplementary table S4-1: Performance measures of the XGBoost single- and multi-omics prediction models
  - Supplementary table S4-2: Features and mean gain (importance) of the XGBoost single- and multi-omics prediction models
  - Supplementary table S4-3: Distribution of booster-types observed in XGBoost models
- **Table S5:** Results of linear mixed models for metabolic parameters, related to Fig. 4, Fig. 5, Fig.6.
- **Table S6** Compliance markers (anthropometrics and resting energy expenditure) before and after intervention, related to compliance section in STAR methods
- **Table S7:** provided in separate excel file, related to STAR methods
  - Supplementary table S7-1: Performance characteristics of DIABLO models with different design settings to characterize RS and NRS at baseline
  - Supplementary table S7-2: Performance characteristics of DIABLO models with different design settings to characterize the change in multi-omics features of RS and NRS
  - Supplementary table S7-3: Comparing the BER of different distance metrics (max, mahalanobis, centroids) for the best DIABLO models characterizing RS and NRS at baseline
  - Supplementary table S7-4: Comparing the BER of different distance metrics (max, mahalanobis, centroids) for the best DIABLO models characterizing the change in multi-omics features of RS and NRS

**Figures:**

- **Figure S1:** CONSORT flow diagram of participants, related to Fig. 1
- **Figure S2:** Top Features from XGBoost prediction models, related to Fig. 2 and Fig. 3
- **Figure S3:** Overview of data pre-processing and models, related to data preparation section in STAR methods

**Table S2: Additional baseline characteristics of RS, NRS and all participants in the different diet groups, related to Fig. 2 and Fig. 3.**

| **Parameters** | | **All diets combined** | | | | **HD** | | | | **ND** | | | | **VD** | | | |
| --- | --- | --- | --- | --- | --- | --- | --- | --- | --- | --- | --- | --- | --- | --- | --- | --- | --- |
|  |  | **All**  *n*=  138 | **RS**  *n*=46 | **NRS**  *n*=92 | ***p*-value**  **(*p*_adj_^3^)** | **All**  *n*=46 | **RS**  *n*=10 | **NRS**  *n*=36 | ***p*-value**  **(*p*_adj_^3^)** | **All**  *n*=47 | **RS**  *n*=27 | **NRS**  *n*=20 | ***p*-value**  **(*p*_adj_^3^)** | **All**  *n*=45 | **RS**  *n*=9 | **NRS**  *n*=36 | ***p*-value**  **(*p*_adj_^3^)** |
| **General** | Sex  (women/men. %)^1^ | 59/41 | 70/30 | 54/46 | 0.086 | 52/48 | 50/50 | 53/47 | 0.876 | 66/34 | 74/26 | 55/45 | 0.172 | 60/40 | 78/22 | 56/44 | 0.224 |
|  | Age  (years) | 59±1 | 60±1 | 59±1 | 0.895 | 59±1 | 58±2 | 59±1 | 0.611 | 60±1 | 60±1 | 61±2 | 0.581 | 59±1 | 61±2 | 59±1 | 0.464 |
| **Anthro-pometry & blood pressure** | BMI  (kg/m^2^) | 31  ±0.3 | 30.9  ±0.5 | 31.1  ±0.4 | 0.736 (>.999) | 31.1  ±0.5 | 30.8  ±1.1 | 31.2  ±0.6 | 0.779  (>.999) | 30.7  ±0.5 | 30.6  ±0.6 | 30.9  ±0.9 | 0.725  (>.999) | 31.3  ±0.6 | 32  ±1 | 31.1  ±0.7 | 0.564  (>.999) |
|  | Waist-to-height ratio | 0.62  ±0 | 0.63  ±0 | 0.61  ±0 | 0.161 (>.999) | 0.61  ±0 | 0.6  ±0 | 0.61  ±0 | 0.442  (>.999) | 0.62  ±0 | 0.63  ±0 | 0.6  ±0 | 0.054  (0.432) | 0.63  ±0 | 0.65  ±0 | 0.62  ±0 | 0.188  (>.999) |
|  | Waist-to-hip ratio | 0.96±0.01 | 0.96±0.01 | 0.96±0.01 | 0.981  (>.999) | 0.96±0.01 | 0.97  ±0.03 | 0.96  ±0.01 | 0.743  (>.999) | 0.95  ±0.01 | 0.95  ±0.01 | 0.95  ±0.02 | 0.88  (>.999) | 0.95  ±0.01 | 0.95  ±0.01 | 0.96  ±0.01 | 0.781  (>.999) |
|  | Fat mass  (%) | 41.3  ±0.7 | 42.4  ±1.1 | 40.8  ±0.9 | 0.285 (>.999) | 40  ±1.3 | 39  ±2.6 | 40.3  ±1.5 | 0.696  (>.999) | 41.3  ±1.2 | 41.9  ±1.4 | 40.6  ±2 | 0.565  (>.999) | 42.6  ±1.2 | 47.3  ±1.5 | 41.4  ±1.4 | **0.046**  (0.322) |
|  | SBP  (mmHg) | 136±1 | 135±3 | 136±2 | 0.744 (>.999) | 134±2 | 127±3 | 136±3 | **0.035**  (0.28) | 136±2 | 137±4 | 135±3 | 0.679  (>.999) | 138±3 | 140±9 | 137±3 | 0.768  (>.999) |
|  | DBP  (mmHg) | 87±1 | 88±2 | 87±1 | 0.527 (>.999) | 86±2 | 82±3 | 88±2 | 0.091  (0.637) | 88±2 | 90±2 | 85±2 | 0.11  (0.77) | 88±2 | 91±4 | 88±2 | 0.377  (>.999) |
|  | Pulse  (bpm) | 68±1 | 69±2 | 67±1 | 0.563 (>.999) | 68±2 | 67±3 | 69±2 | 0.547  (>.999) | 67±2 | 67±2 | 67±2 | 0.997  (>.999) | 68±2 | 75±3 | 66±2 | **0.02**  (0.16) |
|  | RMR  (kcal/day) | 1713  ±25 | 1682  ±39 | 1729  ±31 | 0.371 (>.999) | 1784  ±43 | 1821  ±98 | 1773  ±48 | 0.65  (>.999) | 1683  ±39 | 1658  ±51 | 1716  ±62 | 0.464  (>.999) | 1673  ±45 | 1599  ±57 | 1691  ±54 | 0.42  (>.999) |
| **Lipid metabolism** | TC  (mmol/l) | 5.6  ±0.1 | 6  ±0.2 | 5.4  ±0.1 | **0**  (**<0.001**) | 5.3  ±0.2 | 5.7  ±0.3 | 5.1  ±0.2 | 0.1  (0.4) | 6  ±0.2 | 6.1  ±0.2 | 5.7  ±0.2 | 0.152  (0.65) | 5.6  ±0.1 | 6.1  ±0.4 | 5.4  ±0.1 | 0.058  (0.232) |
|  | LDL-C  (mmol/l) | 3.8  ±0.1 | 4.3  ±0.2 | 3.6  ±0.1 | **0**  (**<0.001**) | 3.5  ±0.1 | 4  ±0.4 | 3.4  ±0.1 | 0.07  (0.35) | 4.1  ±0.1 | 4.3  ±0.2 | 3.9  ±0.2 | 0.16  (0.65) | 3.8  ±0.1 | 4.5  ±0.4 | 3.7  ±0.1 | **0.012**  (0.06) |
|  | HDL-C  (mmol/l) | 1.5  ±0 | 1.6  ±0.1 | 1.4  ±0 | **0.036**  **(**0.108) | 1.5  ±0.1 | 1.7  ±0.1 | 1.4  ±0.1 | 0.121  (0.4) | 1.5  ±0.1 | 1.6  ±0.1 | 1.4  ±0.1 | 0.13  (0.65) | 1.4  ±0.1 | 1.4  ±0.1 | 1.4  ±0.1 | 0.929  (>.999) |
|  | Total fasting TAG (mmol/l) | 1.6  ±0.1 | 1.5  ±0.1 | 1.6  ±0.1 | 0.336 (0.672) | 1.4  ±0.1 | 1.3  ±0.1 | 1.4  ±0.1 | 0.604  (0.608) | 1.6  ±0.1 | 1.5  ±0.1 | 1.7  ±0.1 | 0.28  (0.65) | 1.7  ±0.1 | 1.7  ±0.1 | 1.7  ±0.1 | 0.911  (>.999) |
|  | NEFA  (mmol/l) | 0.54  ±0.02 | 0.56  ±0.03 | 0.54  ±0.02 | 0.611 (0.672) | 0.5  ±0.03 | 0.45  ±0.06 | 0.51  ±0.03 | 0.304  (0.608) | 0.55  ±0.03 | 0.57  ±0.04 | 0.52  ±0.03 | 0.391  (0.65) | 0.58  ±0.03 | 0.63  ±0.06 | 0.57  ±0.04 | 0.459  (>.999) |
| **Glucose & insulin metabolism** | Glucose  (mmol/l) | 5.2  ±0.1 | 5.3  ±0.1 | 5.2  ±0.1 | 0.285 (>.999) | 5.2  ±0.1 | 5.3  ±0.2 | 5.2  ±0.1 | 0.597  (>.999) | 5.2  ±0.1 | 5.2  ±0.1 | 5.1  ±0.1 | 0.407  (>.999) | 5.1  ±0.1 | 5.2  ±0.3 | 5.1  ±0.1 | 0.508  (>.999) |
|  | Insulin  (pmol/l) | 94  ±4.8 | 87.1  ±6.4 | 97.5  ±6.4 | 0.308 (>.999) | 93.8  ±9 | 68.6  ±11.5 | 100.8  ±10.8 | 0.14  (>.999) | 85.4  ±6 | 86.8  ±7.6 | 83.4  ±9.9 | 0.78  (>.999) | 103.3  ±9.6 | 108.5  ±18.3 | 102  ±11.2 | 0.793  (>.999) |
|  | HOMA-IR | 3.1  ±0.2 | 2.9  ±0.2 | 3.2  ±0.2 | 0.424 (>.999) | 3.1  ±0.3 | 2.3  ±0.4 | 3.3  ±0.4 | 0.175  (>.999) | 2.8  ±0.2 | 2.9  ±0.3 | 2.6  ±0.3 | 0.612  (>.999) | 3.3  ±0.3 | 3.6  ±0.7 | 3.3  ±0.4 | 0.689  (>.999) |
|  | HbA1c  (%) | 5.4  ±0 | 5.4  ±0.1 | 5.4  ±0 | 0.546 (>.999) | 5.5  ±0.1 | 5.4  ±0.1 | 5.5  ±0.1 | 0.805  (>.999) | 5.4±  0.1 | 5.4  ±0.1 | 5.4±  0.1 | 0.797  (>.999) | 5.4±  0.1 | 5.5  ±0.1 | 5.3  ±0.1 | 0.267  (>.999) |
|  | Matsuda Index | 3.7  ±0.2 | 3.7  ±0.3 | 3.8  ±0.2 | 0.86 (>.999) | 4.1  ±0.4 | 5.2  ±0.9 | 3.8  ±0.4 | 0.12  (>.999) | 3.7  ±0.2 | 3.4  ±0.2 | 4.2  ±0.5 | 0.089  (0.623) | 3.4  ±0.3 | 3  ±0.6 | 3.5  ±0.4 | 0.507  (>.999) |
|  | Insulinogenic Index^2^ | 2  ±0.3 | 1.3  ±0.2 | 2.4  ±0.5 | **0.007** (0.7) | 2.8  ±0.9 | 0.9  ±0.2 | 3.3  ±1.1 | **0.038**  (0.38) | 1.5  ±0.2 | 1.1  ±0.1 | 1.9  ±0.3 | 0.051  (0.408) | 1.7  ±0.3 | 2.1  ±0.6 | 1.6  ±0.4 | 0.665  (>.999) |
|  | Disposition Index^2^ | 6.3  ±1.1 | 4.3  ±0.7 | 7.3  ±1.6 | 0.061 (0.549) | 9  ±2.9 | 3.9  ±0.5 | 10.5  ±3.7 | 0.783  (>.999) | 5.5  ±0.8 | 3.6  ±0.4 | 8  ±1.8 | **0.01**  (0.1) | 4.3  ±1.3 | 6.9  ±3.2 | 3.7  ±1.4 | 0.791  (>.999) |
|  | OGIS | 361  ±7 | 350  ±12 | 366  ±9 | 0.326 (>.999) | 363  ±15 | 396  ±27 | 354  ±17 | 0.245  (>.999) | 374  ±11 | 349  ±15 | 407  ±14 | **0.01**  (0.1) | 345  ±12 | 305  ±24 | 355  ±14 | 0.109  (>.999) |
|  | QUICKI | 0.33  ±0 | 0.33  ±0 | 0.33  ±0 | 0.686 (>.999) | 0.34  ±0 | 0.35  ±0.01 | 0.33  ±0.01 | 0.174  (>.999) | 0.34  ±0 | 0.33  ±0 | 0.34  ±0.01 | 0.5  (>.999) | 0.33  ±0 | 0.33  ±0.01 | 0.33  ±0 | 0.607  (>.999) |
|  | Fasting GLP‑1  (pmol/l) | 3.8  ±0.1 | 4.1  ±0.3 | 3.7  ±0.1 | 0.208 (>.999) | 3.7  ±0.2 | 4.1  ±0.6 | 3.6  ±0.2 | 0.376  (>.999) | 4  ±0.3 | 4.4  ±0.4 | 3.5  ±0.3 | 0.122  (0.732) | 3.7  ±0.2 | 3.3±  0.5 | 3.9  ±0.2 | 0.283  (>.999) |
| **Kidney function** | Creatinine (µmol/L) | 70.9  ±1.3 | 68.7  ±2.2 | 72.1  ±1.6 | 0.227 (0.908) | 69.3  ±2 | 70.9  ±3.4 | 68.9  ±2.4 | 0.682  (>.999) | 70.7  ±2.4 | 67.6  ±3.3 | 74.9  ±3.4 | 0.142  (0.404) | 72.8  ±2.4 | 69.4  ±3.5 | 73.7  ±2.9 | 0.487  (>.999) |
|  | Urea  (mmol/L) | 5.1  ±0.1 | 5.2  ±0.2 | 5  ±0.1 | 0.299 (0.908) | 4.9  ±0.2 | 4.6  ±0.4 | 5  ±0.2 | 0.339  (>.999) | 5.2  ±0.2 | 5.5  ±0.2 | 4.9  ±0.3 | 0.128  (0.404) | 5  ±0.2 | 5.2  ±0.5 | 5  ±0.2 | 0.599  (>.999) |
|  | Uric acid  (µmol/L) | 322  ±0 | 315  ±10 | 325  ±7 | 0.378 (0.908) | 321  ±10 | 298  ±16 | 327  ±11 | 0.216  (0.864) | 317  ±9 | 304  ±13 | 334  ±12 | 0.101  (0.404) | 327  ±10 | 366  ±20 | 318  ±11 | **0.046**  **(**0.184) |
|  | GFR (mL/min) | 93.1  ±1.1 | 94  ±1.8 | 92.6  ±1.5 | 0.57 (0.908) | 95.7  ±1.7 | 96.1  ±2.9 | 95.6  ±2.1 | 0.905  (>.999) | 91.6  ±1.9 | 93.8  ±2.4 | 88.5  ±2.8 | 0.16  (0.404) | 91.9  ±2.3 | 92.1  ±4.8 | 91.9  ±2.6 | 0.976  (>.999) |
| **SCFA** | Acetic acid stool (µmol/g) | 37.9  ±2.3 | 38.7  ±3.7 | 37.5  ±2.9 | 0.798 (>.999) | 40.5  ±4.3 | 51.5  ±9.7 | 37.3  ±4.7 | 0.169  0.507) | 36.4  ±3.2 | 36  ±4.3 | 37.2  ±4.9 | 0.849  (>.999) | 36.8  ±4.4 | 32.9  ±7.6 | 37.9  ±5.1 | 0.651  (>.999) |
|  | Butyric acid stool (µmol/g) | 91.3  ±2.6 | 89.7  ±4 | 92.2  ±3.4 | 0.648 (>.999) | 98.1  ±5 | 104.6  ±11.5 | 96.1  ±5.5 | 0.482  (0.964) | 85.2  ±3.5 | 84.7  ±4.5 | 86  ±5.7 | 0.865  (>.999) | 90.6  ±5 | 88  ±7.6 | 91.3  ±5.6 | 0.778  (>.999) |
| **Liver function** | GGT  (U/L) | 32±2 | 27±2 | 34±3 | 0.058 (0.232) | 28±2 | 23±3 | 30±3 | 0.211  (0.636) | 31±3 | 28±2 | 35±7 | 0.384  (>.999) | 36±4 | 30±6 | 37±5 | 0.509  (>.999) |
|  | ALT  (U/L) | 29±1 | 28±2 | 30±1 | 0.375 (>.999) | 27±2 | 23±3 | 29±2 | 0.159  (0.636) | 28±2 | 28±2 | 28±3 | 0.964  (>.999) | 32±2 | 32±7 | 31±2 | 0.876  (>.999) |
|  | AST  (U/L) | 27±1 | 26±1 | 28±1 | 0.36 (>.999) | 26±1 | 26±2 | 26±2 | 0.922  (>.999) | 27±1 | 26±1 | 29±3 | 0.281  (>.999) | 27±1 | 26±2 | 28±1 | 0.542  (>.999) |
|  | Bilirubin  (µmol/L) | 10  ±0.4 | 9.2  ±0.7 | 9.8  ±0.4 | 0.487 (>.999) | 9  ±0.7 | 9.3  ±2.1 | 8.9  ±0.7 | 0.839  (>.999) | 9.5  ±0.6 | 9.4  ±0.9 | 9.7  ±0.7 | 0.804  (>.999) | 10.2  ±0.6 | 8.6  ±0.6 | 10.6  ±0.8 | 0.194  (0.776) |
| **PGS** | PGS_LDL-C_ | ‑0.04±0.99 | 0.18  ±0.15 | ‑0.17±0.11 | 0.059 (0.236) | ‑0.23±1.1 | 0  ±0.29 | -0.31  ±0.2 | 0.446  (>.999) | 0.25  ±0.96 | 0.5  ±0.18 | ‑0.13  ±0.24 | **0.038**  (0.152) | ‑0.14  ±0.84 | -0.5  ±0.35 | ‑0.05  ±0.13 | 0.152  (0.456) |
|  | PGS_HDL-C_ | 0.02  ±0.99 | 0  ±0.15 | 0.02  ±0.11 | 0.911 (>.999) | 0.14  ±0.93 | 0.07  ±0.3 | 0.16  ±0.17 | 0.799  (>.999) | -0.2  ±1.01 | ‑0.17  ±0.21 | ‑0.23  ±0.25 | 0.854  (0.854) | 0.1  ±1.02 | 0.41  ±0.31 | 0.01  ±0.18 | 0.307  (0.614) |
|  | PGS_TC_ | ‑0.01±0.97 | 0.14  ±0.14 | ‑0.08±0.11 | 0.224 (0.672) | ‑0.1  ±1.16 | 0.11  ±0.32 | ‑0.17  ±0.21 | 0.509  (>.999) | 0.19  ±0.97 | 0.38  ±0.18 | -0.1  ±0.26 | 0.117  (0.351) | ‑0.11  ±0.74 | ‑0.52  ±0.25 | 0.01  ±0.12 | 0.058  (0.232) |
|  | PGS_TAG_ | ‑0.08±1.06 | ‑0.04±0.16 | ‑0.09±0.12 | 0.801 (>.999) | ‑0.12±1.09 | 0.33  ±0.27 | -0.26  ±0.2 | 0.139  (0.556) | ‑0.01  ±1.04 | ‑0.17  ±0.22 | 0.25  ±0.22 | 0.201  (0.402) | -0.1  ±1.08 | -0.1  ±0.41 | -0.1  ±0.18 | 0.992  (0.992) |
| **Inflam-mation & gut integrity** | hsCRP  (mg/l) | 2.9  ±0.2 | 3  ±0.4 | 2.9  ±0.3 | 0.951  (>.999) | 3.1  ±0.4 | 2.6  ±0.8 | 3.2  ±0.5 | 0.558  (>.999) | 2.9  ±0.4 | 3.2  ±0.5 | 2.6  ±0.5 | 0.416  (0.832) | 2.8  ±0.4 | 2.6  ±0.6 | 2.8  ±0.5 | 0.845  (0.845) |
|  | LPS  (pg/ml) | 15.7  ±1.8 | 14  ±2.3 | 16.5  ±2.5 | 0.522  (>.999) | 18.9  ±3.9 | 20.4  ±7.4 | 18.4  ±4.6 | 0.842  (>.999) | 11.6  ±1.7 | 13.3  ±2.6 | 9.3  ±2.1 | 0.266  (0.798) | 16.7  ±3.5 | 9.1  ±2.9 | 18.5  ±4.2 | 0.281  (0.562) |
|  | Zonulin^2^  (ng/mL) | 297  ±21 | 326  ±36 | 282  ±26 | 0.205 (0.615) | 279  ±40 | 268  ±103 | 282  ±44 | 0.627  (>.999) | 292  ±34 | 303  ±42 | 278  ±59 | 0.423  (0.832) | 320  ±36 | 463  ±64 | 284  ±40 | **0.017**  (0.051) |
| **Postprandial**  **values** | Postprandial glucose (mmol/L*min) | 3167±17 | 334±29 | 308±20 | 0.473 (>.999) | 311±27 | 255  ±3 | 326  ±33 | 0.111  (0.444) | 300  ±30 | 331  ±36 | 258  ±50 | 0.225  (0.832) | 340  ±31 | 428  ±97 | 318  ±30 | 0.156  (0.624) |
|  | Postprandial insulin (pmol/L*min) | 101741  ±5360 | 96419±8588 | 104402  ±6813 | 0.485 (>.999) | 96419±9318 | 62547  ±11178 | 105827  ±11049 | 0.054  (0.27) | 88817  ±5898 | 96323  ±7813 | 78684  ±8703 | 0.141  (0.705) | 120680  ±11525 | 134346  ±32092 | 117264  ±12175 | 0.559  (>.999) |
|  | Total postprandial TAG (mmol/L*min) | 183.6±6.6 | 174.1±9.5 | 188.8±8.7 | 0.3 (>.999) | 169.5±10.7 | 162.9  ±17.3 | 171.3  ±12.9 | 0.751  (0.89) | 181.5  ±10.1 | 170.4  ±13 | 196.4  ±15.8 | 0.208  (0.832) | 201.8  ±13.3 | 200.6  ±22.4 | 202  ±15.6 | 0.968  (>.999) |
|  | Postprandial NEFA (mmol/L*min) | 31.85±0.94 | 33.17±1.85 | 31.18±1.06 | 0.354 (>.999) | 29.32±1.5 | 25.99  ±3.08 | 30.24  ±1.71 | 0.247  (0.741) | 32.61  ±1.52 | 33.18  ±2.34 | 31.83  ±1.74 | 0.645  (>.999) | 33.74  ±1.82 | 41.1  ±4.21 | 31.79  ±1.91 | 0.036  (0.19) |
|  | Postprandial GLP‑1 (pmol/L*min) | 1245  ±73 | 1261  ±128 | 1237  ±89 | 0.874 (>.999) | 1161  ±118 | 1334  ±325 | 1112  ±123 | 0.445  (0.89) | 1369  ±149 | 1285  ±177 | 1481  ±259 | 0.522  (>.999) | 1202  ±106 | 1109  ±157 | 1225.  ±128 | 0.666  (>.999) |

*Mean + SEM. p-value from student’s t-test unless stated otherwise. ^1^p-value from Pearson’s chi-square test. ^2^p-value from Mann-Whitney U Test of log transformed data. ^3^p-value adjusted block-wise for multiple testing by Bonferroni-Holm correction *p≤0.05; **p≤0.01; ***p≤0.001 ;****p≤0.0001. Results for “all diets combined” were confirmed by sensitivity analysis.*

*ALT: alanine transaminase; AST: aspartate transferase; BMI: body mass Index; DBP: diastolic blood pressure; GFR: glomerular filtration rate; GGT: gamma-glutamyl transferase; GLP-1: glucagon like peptide 1; HbA1c: hemoglobin A1c; HD: Habitual Diet; HDL-C: high density lipoprotein cholesterol; HOMA-IR: homeostasis model assessment for insulin resistance; hsCRP: high sensitivity C-Reactive Protein; LDL-C: low density lipoprotein cholesterol; LPS: lipopolysaccharide; ND: Nordic Diet; NEFA: non-esterified fatty acids; NRS: Non-Responders; OGIS: oral glucose insulin sensitivity index; PGS: polygenetic risk score; QUICKI: quantitative insulin sensitivity check index; RMR: resting metabolic rate; RS: Responders SBP: systolic blood pressure; TAG: triacylglycerides; TC: total cholesterol; VD: Vegetarian Diet.*

**Table S5: Results of linear mixed models for metabolic parameters, related to Fig. 4, Fig. 5, Fig.6.**

|  |  | **All diets combined**  ***n*=138** | | **HD** | | **ND** | | **VD** | |
| --- | --- | --- | --- | --- | --- | --- | --- | --- | --- |
|  |  | p_unadj_ | p_adj_ | p_unadj_ | p_adj_ | p_unadj_ | p_adj_ | p_unadj_ | p_adj_ |
| **Anthropometry & blood pressure** | BMI (kg/m^2^)^1^ | 0.898 | > .999 | 0.495 | > .999 | 0.552 | > .999 | 0.582 | > .999 |
|  | Waist-to-height ratio | 0.898 | > .999 | 0.207 | > .999 | 0.586 | > .999 | 0.530 | > .999 |
|  | Waist-to-hip ratio^1^ | 0.376 | > .999 | 0.720 | > .999 | 0.533 | > .999 | 0.078 | 0.624 |
|  | Fat mass (%) | 0.354 | > .999 | 0.908 | > .999 | 0.831 | > .999 | 0.742 | > .999 |
|  | SBP mmHg) | 0.797 | > .999 | 0.672 | > .999 | 0.924 | > .999 | 0.330 | > .999 |
|  | DBP (mmHg) | 0.869 | > .999 | 0.735 | > .999 | 0.923 | > .999 | 0.503 | > .999 |
|  | Pulse (bpm) | 0.582 | > .999 | 0.098 | 0.784 | 0.126 | > .999 | 0.432 | > .999 |
|  | RMR (kcal/day) | 0.099 | 0.792 | 0.849 | > .999 | 0.170 | > .999 | 0.816 | > .999 |
| **Lipid metabolism** | TC (mmol/l)^1^ | **0.000** | **0.000** | **0.000** | **0.000** | **0.000** | **0.000** | **0.000** | **0.000** |
|  | LDL-C (mmol/l)^1^ | **0.000** | **0.000** | **0.000** | **0.000** | **0.000** | **0.000** | **0.000** | **0.000** |
|  | HDL-C (mmol/l)^1^ | **0.000** | **0.000** | **0.008** | **0.024** | **0.014** | **0.042** | **0.047** | 0.141 |
|  | Total fasting TAG (mmol/l)^1^ | 0.639 | 0.639 | **0.040** | 0.08 | 0.588 | > .999 | 0.211 | 0.211 |
|  | NEFA (mmol/l) | 0.203 | 0.400 | 0.597 | 0.597 | 0.756 | > .999 | **0.047** | 0.141 |
| **Glucose & insulin metabolism** | Glucose (mmol/l)^1^ | 0.130 | > .999 | 0.385 | > .999 | 0.364 | > .999 | 0.884 | > .999 |
|  | Insulin (pmol/l) | 0.397 | > .999 | 0.950 | > .999 | 0.309 | > .999 | 0.579 | > .999 |
|  | HOMA-IR | 0.340 | > .999 | 0.916 | > .999 | 0.199 | > .999 | 0.485 | > .999 |
|  | HbA1c (%)^1^ | 0.908 | > .999 | 0.591 | > .999 | 0.900 | > .999 | 0.844 | > .999 |
|  | Matsuda Index | 0.153 | > .999 | 0.704 | > .999 | 0.061 | 0.61 | 0.680 | > .999 |
|  | Insulinogenic Index | 0.428 | > .999 | 0.646 | > .999 | 0.516 | > .999 | 0.470 | > .999 |
|  | Disposition Index | 0.168 | > .999 | 0.686 | > .999 | 0.421 | > .999 | 0.128 | > .999 |
|  | OGIS | 0.716 | > .999 | 0.745 | > .999 | 0.652 | > .999 | 0.908 | > .999 |
|  | QUICKI^1^ | 0.246 | > .999 | 0.947 | > .999 | 0.422 | > .999 | 0.891 | > .999 |
|  | Fasting GLP‑1 (pmol/l) | 0.916 | > .999 | 0.391 | > .999 | 0.439 | > .999 | **0.019** | 0.19 |
| **Kidney function** | Creatinine (µmol/L) | 0.214 | 0.642 | 0.689 | > .999 | 0.900 | > .999 | 0.275 | 0.825 |
|  | Urea (mmol/L) | 0.341 | 0.642 | 0.297 | 0.891 | 0.385 | > .999 | 0.411 | 0.825 |
|  | Uric acid (µmol/L) | 0.136 | 0.544 | 0.051 | 0.204 | 0.787 | > .999 | 0.390 | 0.825 |
|  | GFR (mL/min) | 0.226 | 0.642 | 0.627 | > .999 | 0.937 | > .999 | 0.126 | 0.504 |
| **Liver function** | GGT (U/L) | 0.605 | > .999 | 0.895 | > .999 | 0.792 | > .999 | 0.554 | > .999 |
|  | ALT (U/L) | 0.264 | 0.792 | 0.208 | 0.624 | 0.234 | 0.936 | 0.803 | > .999 |
|  | AST (U/L) | 0.178 | 0.712 | **0.018** | 0.072 | 0.593 | > .999 | 0.790 | > .999 |
|  | Bilirubin (µmol/L) | 0.935 | > .999 | 0.746 | > .999 | 0.935 | > .999 | 0.765 | > .999 |
| **SCFA** | Acetic acid stool (µmol/g) | 0.787 | > .999 | 0.295 | 0.885 | 0.505 | > .999 | 0.938 | > .999 |
|  | Butyric acid stool (µmol/g) | 0.103 | 0.309 | 0.897 | > .999 | 0.198 | 0.594 | 0.761 | > .999 |
| **Inflammation & gut integrity** | hsCRP (mg/l) | 0.933 | > .999 | 0.582 | > .999 | 0.536 | > .999 | 0.521 | 0.521 |
|  | LPS (pg/ml) | 0.608 | > .999 | 0.793 | > .999 | 0.477 | > .999 | 0.077 | 0.231 |
|  | Zonulin (ng/mL) | 0.514 | > .999 | 0.908 | > .999 | 0.977 | > .999 | 0.221 | 0.442 |
| **Postprandial values** | Postprandial glucose (mmol/L*min) | 0.718 | > .999 | 0.277 | > .999 | 0.273 | 0.819 | 0.870 | > .999 |
|  | Postprandial insulin (pmol/L*min) | 0.115 | 0.575 | 0.922 | > .999 | 0.068 | 0.34 | 0.557 | > .999 |
|  | Total postprandial TAG (mmol/L*min) | 0.713 | > .999 | 0.052 | 0.26 | 0.184 | 0.736 | 0.517 | > .999 |
|  | Postprandial NEFA (mmol/L*min) | 0.828 | > .999 | 0.527 | > .999 | 0.811 | 0.819 | 0.068 | 0.34 |
|  | Postprandial GLP‑1 (pmol/L*min) | 0.650 | > .999 | 0.434 | > .999 | 0.321 | 0.819 | 0.393 | > .999 |

*Compared using linear mixed effect models; p-value refers to the null hypothesis that the change in outcome parameters over time (visits) is identical across all Responder status and diet groups (interaction term of group × status x visit). A study variable is included in all models. p_unadj_: raw p-value; p_adj_: adjusted block-wise for multiple testing by Bonferroni-Holm correction *p≤0.05; **p≤0.01; ***p≤0.001 ;****p≤0.0001.*

*ALT: alanine transaminase; AST: aspartate transferase; BMI: body mass Index; DBP: diastolic blood pressure; GFR: glomerular filtration rate; GGT: gamma-glutamyl transferase; GLP-1: glucagon like peptide 1; HbA1c: hemoglobin A1c; HD: Habitual Diet; HDL-C: high density lipoprotein cholesterol; HOMA-IR: homeostasis model assessment for insulin resistance; hsCRP: high sensitivity C-Reactive Protein; LDL-C: low density lipoprotein cholesterol; LPS: lipopolysaccharide; ND: Nordic Diet; NEFA: non-esterified fatty acids; NRS: Non-Responders; OGIS: oral glucose insulin sensitivity index; PGS: polygenetic risk score; QUICKI: quantitative insulin sensitivity check index; RMR: resting metabolic rate; RS: Responders SBP: systolic blood pressure; TAG: triacylglycerides; TC: total cholesterol; VD: Vegetarian Diet.*

**Table S6: Compliance markers (anthropometrics and resting energy expenditure) before and after intervention, related to compliance section in STAR methods**

|  | **All diets combined** | | | | | **HD** | | | | | **ND** | | | | | **VD** | | | | |  |
| --- | --- | --- | --- | --- | --- | --- | --- | --- | --- | --- | --- | --- | --- | --- | --- | --- | --- | --- | --- | --- | --- |
|  | **RS**  (*n*=46) | | **NRS**  (*n*=92) | |  | **RS**  (*n*=10) | | **NRS**  (*n*=36) | |  | **RS**  (*n*=27) | | **NRS**  (*n*=20) | |  | **RS**  (*n*=9) | | **NRS**  (*n*=36) | |  |  |
| **Parameters** | **V_B_** | **V_E_** | **V_B_** | **V_E_** | ***p*-value** | **V_B_** | **V_E_** | **V_B_** | **V_E_** | ***p*-value** | **V_B_** | **V_E_** | **V_B_** | **V_E_** | ***p*-value** | **V_B_** | **V_E_** | **V_B_** | **V_E_** | ***p*-value** | ***p*-value global^a^** |
| BMI (kg/m^2^) | 30.8 | 30.6 | 31.5 | 31.2 | 0.9 | 31.2 | 31.2 | 31.7 | 31.5 | 0.5 | 30.2 | 29.8 | 30.4 | 30.1 | 0.6 | 32.2 | 32 | 31.6 | 31.3 | 0.6 | 0.4 |
| Waist-to-height ratio | 0.62 | 0.62 | 0.62 | 0.61 | 0.9 | 0.6 | 0.61 | 0.62 | 0.62 | 0.2 | 0.63 | 0.62 | 0.59 | 0.59 | 0.6 | 0.65 | 0.65 | 0.62 | 0.62 | 0.5 | 0.3 |
| Fat mass (%) | 41.3 | 40.6 | 41.5 | 41.1 | 0.3 | 40.9 | 40.7 | 42.3 | 42.1 | 0.9 | 41.6 | 40.7 | 40.0 | 39.2 | 0.8 | 46.9 | 46.5 | 40.7 | 40.5 | 0.7 | 0.1 |
| RMR (kcal/day) | 1716 | 1642 | 1734 | 1707 | 0.1 | 1774 | 1737 | 1721 | 1692 | 0.9 | 1622 | 1513 | 1666 | 1611 | 0.2 | 1626 | 1614 | 1741 | 1743 | 0.8 | 0.2 |

*Data are shown as mean ± SEM; Compared using linear mixed effect models; p-value refers to the null hypothesis that the change in outcome parameters over time (visits) is identical across all Responder status groups (interaction term of status x visit); ^a^ p-value global refers to the null hypothesis that the change in outcome parameters over time (visits) is identical across all Responder status and diet groups (interaction term of group × status x visit). A study variable is included in all models.*

BMI, body mass index; HD, habitual diet group; ND, Nordic diet group; REE, resting energy expenditure; V_B_, visit baseline; VD, vegetarian diet group; V_E_, visit endline.

**Figure S1: CONSORT flow diagram of participants, related to Fig. 1**

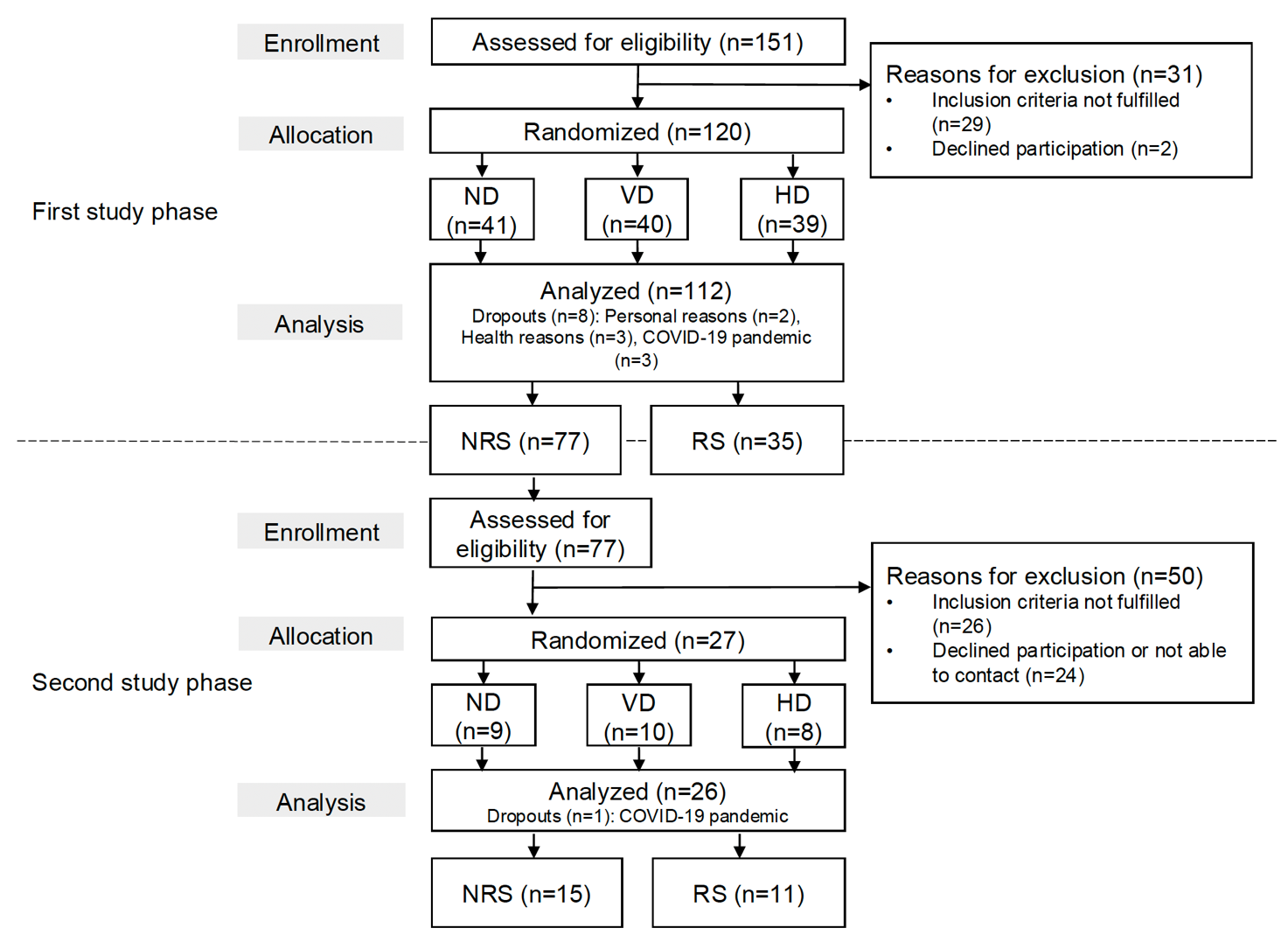

Figure S1 shows the flow diagram of the study participants. For the first study phase, in total 151 individuals were screened for eligibility. 31 persons were excluded for not fulfilling the inclusion criteria or declining participation. 120 participants were randomized into one of three groups: ND, VD or control group (HD). After the intervention of 6-week duration, 39 people completed the study in the ND group, 35 in the VD group and 38 in the HD group. A total of 112 participants completed the first study phase and were subsequently classified into 35 RS and 77 NRS (according to the change in LDL-C during the trial). All 77 NRS were assessed for eligibility for the second study phase which led to an exclusion of 50 participants. Of those, 26 did not fulfill the inclusion criteria (mostly because of a relevant change in their lifestyle after participating in the first study phase such as a reduction in body weight and/or a permanent change to a vegetarian or nordic diet) and 24 participants were either not able to or not interested in participating in the second study phase (due to scheduling overlaps with their current jobs, fear of COVID-19 or personal reasons) or did not respond to the invitation. 27 participants were included and randomized based on their prior diet allocation into one of the two diet groups they had not followed in the first study phase (cross-over-design). 26 of these participants completed the second study phase and were subsequently categorized into 15 NRS and 11 RS.

*CONSORT, consolidated standards of reporting trials; HD, habitual diet; LDL-C, low-density lipoprotein cholesterol; ND, Nordic diet; NRS, Non-Responders; RS, Responders; VD, Vegetarian diet*

**Figure S2: Top features from XGBoost prediction models, related to Fig. 2 and Fig. 3**
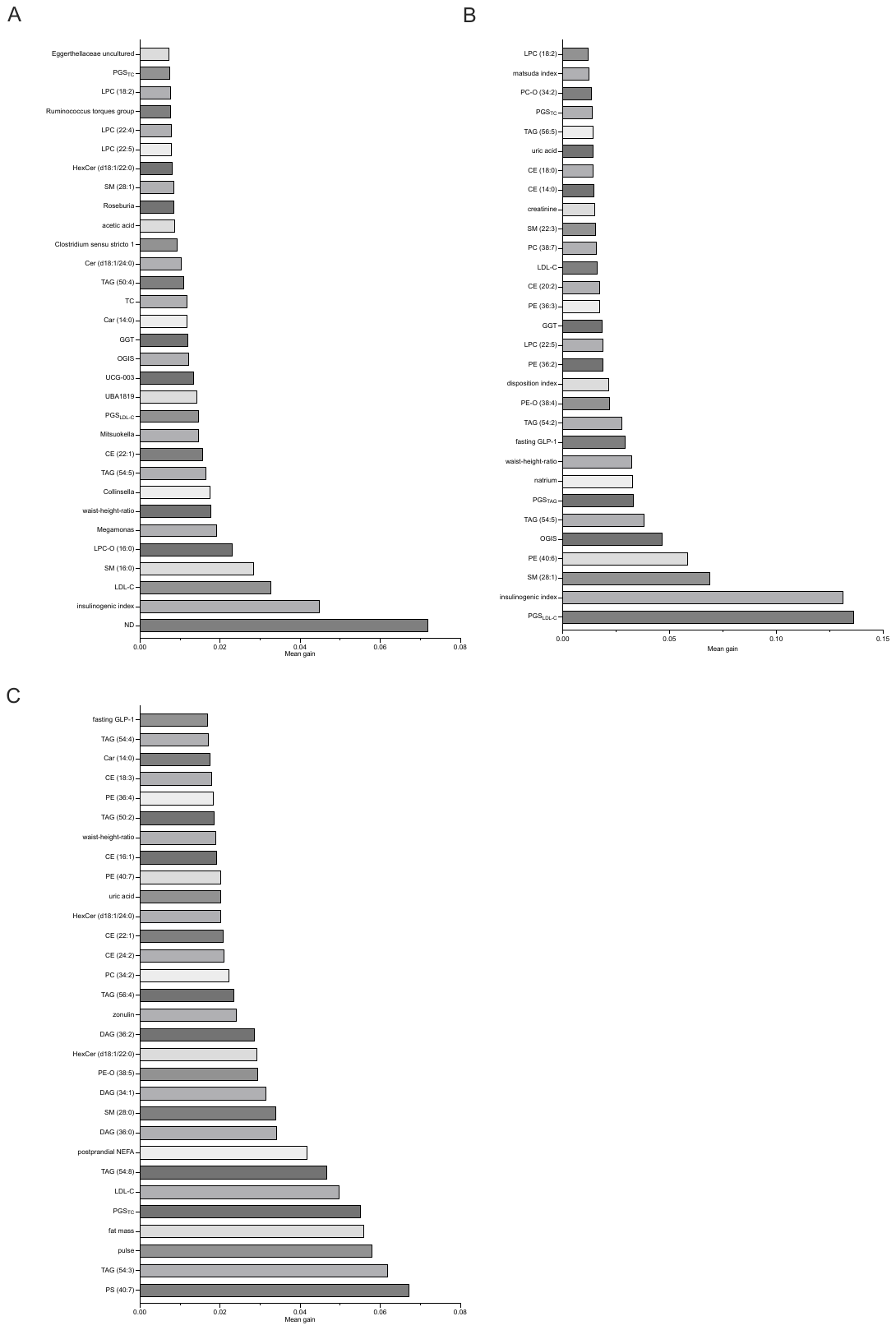

Figures S2 A-C show the 30 features with the highest mean gain values from the XGBpoost prediction models predicting the LDL-C response to the interventions.

A: All diets combined, B: Nordic Diet (ND), C: Vegetarian Diet (VD).

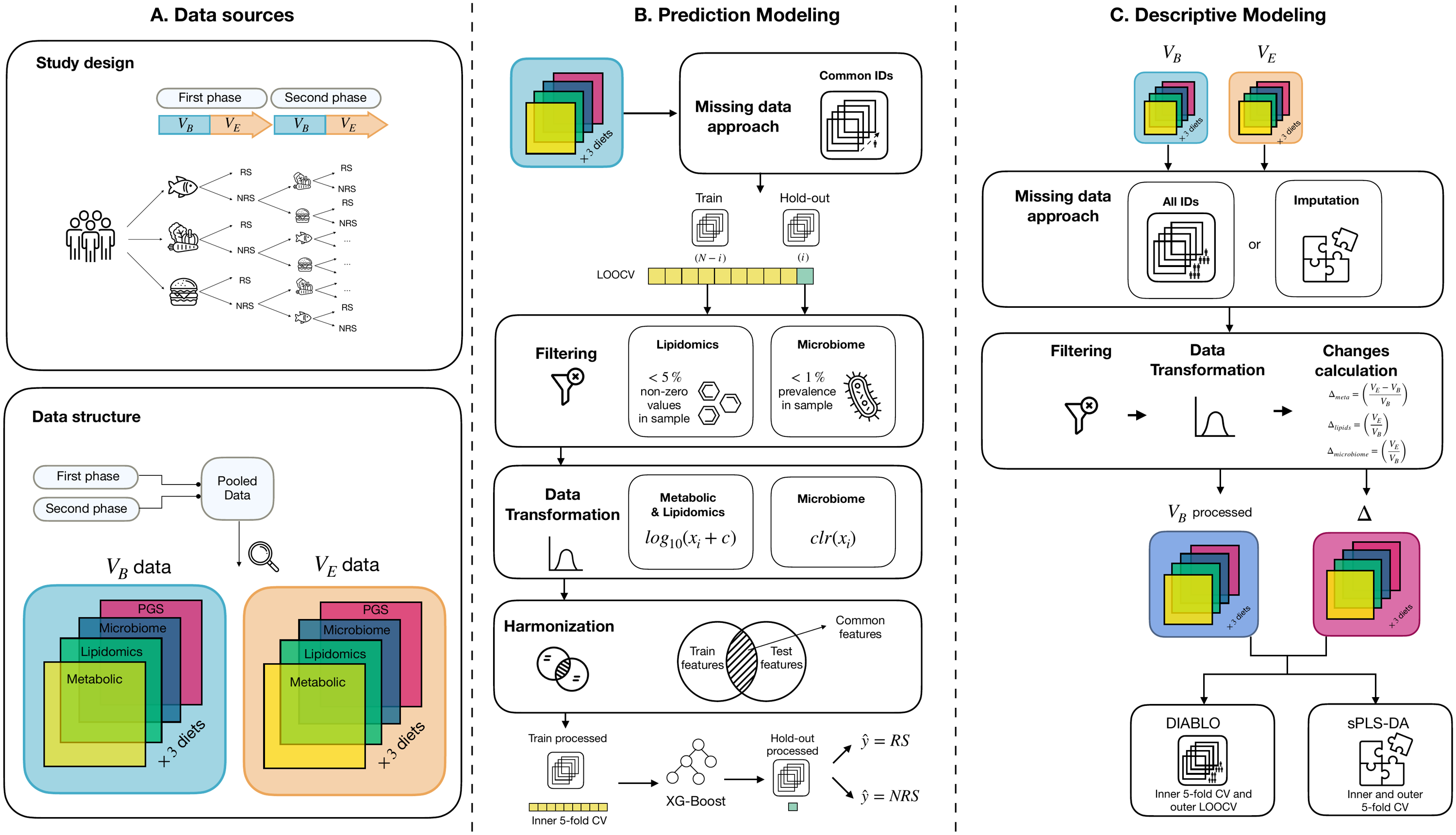
**Figure S3: Overview of data pre-processing and models, related to data preparation section in STAR methods**

Figure S3 shows the pre-processing steps used for preparing datasets for both prediction and descriptive modeling. Panel A shows the study design and the two study phases, in which both baseline and endline omics-datasets were collected. Both study phases were later pooled together into a unique baseline and endline dataset, both containing metabolic, lipidomics, microbiome and PGS data from all participants. Panel B depicts the pre-processing done for the prediction models. Only common participants observed across omics-layers were taken into account for the model. All pre-processing steps were done separately for training and test data following a Leave-One-Out-Cross-Validation (LOOCV) strategy. Specific omics filters and transformations were applied, and features were harmonized across train and test sets. The already processed train data was used to train the model and the hold-out test set was used for prediction. Model performance was evaluated using Sensitivity, Specificity, F1-Score and AUC-ROC metrics. Panel C shows the pre-processing of the datasets for description modeling. All filtering and pre-processing was done analogously to the approach shown in Panel B. Pre-processed baseline and endline data was used for calculating changes across all omics blocks, except for PGS. Descriptive models were then applied using DIABLO and sPLS-DA approaches. This figure was created using designs from www.flaticon.com.
